# Informing individualized multi-scale neural signatures of clozapine response in patients with treatment-refractory schizophrenia

**DOI:** 10.1101/2023.03.10.23286854

**Authors:** Jie Lisa Ji, Todd Lencz, Juan Gallego, Nicholas Neufeld, Aristotle Voineskos, Anil Malhotra, Alan Anticevic

## Abstract

Clozapine is currently the only antipsychotic with demonstrated efficacy in treatment-refractory schizophrenia (TRS). However, response to clozapine differs widely between TRS patients, and there are no available clinical or neural predictive indicators that could be used to increase or accelerate the use of clozapine in patients who stand to benefit. Furthermore, it remains unclear how the neuropharmacology of clozapine contributes to its therapeutic effects. Identifying the mechanisms underlying clozapine’s therapeutic effects across domains of symptomatology could be crucial for development of new optimized therapies for TRS.

Here, we present results from a prospective neuroimaging study that quantitatively related heterogeneous patterns of clinical clozapine response to neural functional connectivity at baseline. We show that we can reliably capture specific dimensions of clozapine clinical response by quantifying the full variation across item-level clinical scales, and that these dimensions can be mapped to neural features that are sensitive to clozapine-induced symptom change. Thus, these features may act as “failure modes” that can provide an early indication of treatment (non-)responsiveness. Lastly, we related the response-relevant neural maps to spatial expression profiles of genes coding for receptors implicated in clozapine’s pharmacology, demonstrating that distinct dimensions of clozapine symptom-informed neural features may be associated with specific receptor targets. Collectively, this study informs prognostic neuro-behavioral measures for clozapine as a more optimal treatment for selected patients with TRS. We provide support for the identification of neuro-behavioral targets linked to pharmacological efficacy that can be further developed to inform optimal early treatment decisions in schizophrenia.

## INTRODUCTION

Schizophrenia is estimated to occur in ∼1% of the population and account for substantial morbidity and mortality worldwide (1). Existing pharmacological treatment options for schizophrenia primarily target dopaminergic receptors (2,3); however, clinical response to dopaminergic drugs can be highly variable and many do not respond. Approximately 30-40% of patients remain refractory with respect to hallucinations and/or delusions and an additional ∼30% of patients experience residual symptoms in other domains, including deficits in mood and cognition (4–6). Indeed, treatment-refractory schizophrenia (TRS) currently accounts for the large majority of health resource utilization associated with psychosis (4). Despite this unmet worldwide clinical burden, treatment options for psychosis spectrum symptoms remain limited.

Currently, clozapine is the only antipsychotic with demonstrable and FDA-indicated efficacy in TRS (7–9). It has also been demonstrated to reduce suicidality, the leading cause of excess mortality in TRS (10). Clozapine is substantially more effective in reducing TRS symptoms, including “positive” symptoms as well as cognitive deficits, compared to all other options (including polypharmacy (11)). However, due to elevated risk of agranulocytosis, patients on clozapine require regular blood monitoring (12). Consequently, clozapine is significantly underutilized in clinical practice (13). Despite its benefits for some TRS patients, a lack of clozapine response is still observed in 40-60% of TRS patients (14), and the time-course of response to clozapine is relatively prolonged (15,16). These findings collectively highlight the heterogeneity of clozapine response, even within patients with TRS. Currently, there are no available clinical or neural predictive indicators that can inform patterns of clozapine response for a given patient (17). Such prognostic biomarkers could increase the utilization of clozapine for patients who are most likely to benefit earlier in their illness course.

In recent years, several promising studies have used functional magnetic resonance imaging (fMRI) methods to derive neural features that are predictive of standard antipsychotic response (18–22). For example, Sarpal et al. have shown that resting-state functional connectivity (rs-FC) of the striatum at baseline can be significantly associated with better future response to 12 weeks of treatment with risperidone or aripiprazole (19). However, no studies to date have examined rs-FC predictors of clozapine response, and only a limited number of fMRI studies have quantified clinical patterns of clozapine response in relation to neural targets (23–26). Therefore, there is a knowledge gap regarding the relationship between clinical clozapine response heterogeneity in TRS and neural targets of such response. In turn, it remains unknown whether patterns of clozapine symptom response can be used to inform individual-specific patterns of neural clozapine response in TRS. Such neural patterns that are indicative of treatment (non-)responsiveness can act as “failure modes” (38), which may inform patient stratification during treatment or enrollment for a clinical trial and thus reduce the risk of failure.

Quantifying the neural targets of clozapine efficacy can inform the identification of underlying mechanisms behind clozapine’s heterogeneous therapeutic effects. This insight is crucial for optimizing new therapies for specific neural targets. For instance, compared to other antipsychotic drugs, clozapine binds to striatal dopamine D2 receptors at lower levels of occupancy (27,28), but at higher levels in cortical and limbic dopamine receptors (29). Additionally, clozapine has a transient effect at D2 receptors, with rapid “fast-off” dissociation that may account for its relatively milder extrapyramidal side effects compared to other first-generation and even second-generation antipsychotics (2,30). Clozapine has also been found to bind to a broad range of other receptors, with significant activity at other dopaminergic, muscarinic, adrenergic, histamine, and serotonergic receptor subtypes (31–33), which have different expression profiles across the brain (34,35). Taken together with the observation that clozapine uptake is widespread across the brain (36,37), different neural targets may underlie different aspects of clozapine response, and thus may present potential clinically-relevant treatment optimization opportunities. However, major knowledge gaps remain in understanding how the neuropharmacology of clozapine contributes to its therapeutic effects.

To address these questions, the current study tested if heterogenous clozapine response patterns in a sample of TRS patients quantitatively relate to alterations in neural functional connectivity prior to clozapine administration. There were two key goals: I) Can we reliably capture individual patterns of clozapine clinical response by quantifying the full variation across item-level clinical scales, as opposed to examining the mean response; II) We tested if rs-FC collected before treatment can inform neural features (38) that are maximally sensitive to patterns of symptoms change. Collectively, we present evidence from a neuroimaging-based clinical study that can inform prognostic neuro-behavioral measures for clozapine as a more optimal treatment for some patients with TRS. We show that individual patterns of clozapine symptom response can be mapped to neural features, which revealed neuro-behavioral targets of clinical efficacy. Lastly, we related response-relevant neural maps to the spatial expression profiles of genes coding for receptors implicated in clozapine’s pharmacology (35). Here we demonstrate that these distinct dimensions of clozapine symptom-informed neural features may be associated with distinct receptor targets. Taken together, these results provide support for the identification of neuro-behavioral pharmacological efficacy targets that can be further developed in the service of optimal early treatment decisions for TRS patients.

## METHODS

### Study Design

This is a 24-week long prospective neuroimaging study in which schizophrenia patients starting clozapine treatment underwent a brain MRI at baseline and then after 12 weeks of treatment. Patients were scanned at two institutions: 1) The Zucker Hillside Hospital (ZHH), the psychiatric hospital for Northwell Health Systems in New York, 2) The Centre for Addiction and Mental Health (CAMH) in Toronto, Canada. Subjects were recruited from the inpatient units, outpatient clozapine clinic, partial hospital programs, and day hospital programs at those institutions. Additionally, subjects were recruited at the New York Presbyterian Hospital – Westchester Division; these subjects were transported to the ZHH site for scanning. The Institutional Review Board at The Zucker Hillside Hospital, the Institutional Review Board at the Centre for Addiction and Mental Health; and the Institutional Review Board at the New York Presbyterian Hospital – Westchester Division gave ethical approval for this work. All subjects were recruited using the same protocol and under the IRB approval obtained from their respective institutions.

#### Inclusion Criteria

1. Age 18-60;
2. DSM-IV diagnosis of schizophrenia and schizoaffective disorder;
3. Patients who are starting clozapine therapy, due to poor responses to two or more antipsychotic drugs with at least 4 weeks for each drug trial;
4. Having moderate to severe psychotic symptoms despite antipsychotic treatment;
5. Able to provide informed consent.

#### Exclusion Criteria

1. Evidence of serious medical conditions,
2. Female patients who are pregnant or breastfeeding;
3. Patients who have failed clozapine therapy in the past;
4. History of allergic reactions to clozapine;
5. Patients who are unable to provide informed consent due to impairment in decision-making ability.
6. Patients who have contraindications to MRI scan
7. Patients who are not appropriate for the research study based on clinical judgment

All subjects were recruited at the start of their clozapine treatment. We sought to recruit patients before treatment initiation but a dose of up to 75 mg per day was allowed for feasibility reasons. Psychopathology ratings included the Brief Psychiatric Rating Scale (BPRS) (39), the Schedule for Assessment of Negative symptoms (SANS), clinical global impression-severity (CGI-S), Calgary Depression Scale for Schizophrenia (CDSS), Young Mania Rating Scale (YMRS), Hillside Hospital Adverse Events Rating Scale (HHAERS), Barnes Rating Scale for Drug-Induced Akathisia (BRSDIA), Simpson-Angus Rating Scale for Extrapyramidal symptoms (SARSES). Adherence to clozapine treatment was established with monthly blood clozapine levels and by patient self report during all research study visits. Clinical rating scales and assessment of medication adherence and substance use were administered during treatment at weeks 1, 2, 3, 4, 6, 8, 10, and 12. Clozapine treatment was managed by the participants’ respective clinicians, and the dose of clozapine was titrated up as patients were able to tolerate. All treating clinicians follow the clozapine treatment guideline set forth by the Zucker Hillside Hospital Pharmacotherapy Committee, which recommends that clozapine treatment should begin with 12.5 mg once or twice daily and then continue with increments of 25 mg every other day until a dose of 100 mg is achieved. After the first 100 mg, clinicians may increase clozapine by 25 to 50 mg/day if well tolerated, to achieve a targeted dose of 300 to 450 mg/day by the end of 2 weeks. Diagnostic interviews were conducted using the Structured Clinical interview for DSM-5 (SCID) and the diagnoses were confirmed after presentation to a panel of experts as part of a consensus conference. Brain MRIs and neuropsychological testing using the MATRICS Consensus Cognitive Battery (MCCB) (40–42) were conducted at baseline and after 12 weeks of treatment. Only baseline scans are considered in this current paper. Demographics information for the sample is shown in **Supplementary Table 1**.

### Neural Data Acquisition

All resting state scans were conducted on a 3T Siemens PRISMA at either the ZHH or CAMH sites; the scanners are identical in technical specifications and have been calibrated by repeated scans of “human phantoms” as previously reported (43). Two 7-minute 17-second resting-state runs were obtained, one each with AP and PA phase encoding directions. Resting scans contained 594 whole-brain volumes, each with 72 contiguous axial/oblique slices in the AC-PC orientation (TR=720ms, TE=33.1ms, matrix = 104x90, FOV = 208mm, voxel = 2x2x2mm, multi-band acceleration factor=8). Of the 30 subjects in the final analysis (see “Data Quality Control” section below), 25 were acquired at ZHH and 5 were acquired at CAMH.

### Symptom Data

The main symptom measures of interest here were the Brief Psychiatric Rating Scale (BPRS, 18 items)(39) for evaluating clinical symptom severity and the MATRICS Consensus Cognitive Battery (MCCB, 7 items)(40–42) for evaluating cognitive performance.

### Dimensionality Reduction of the Symptom Space

For this analysis, we only included those subjects who completed a baseline scan and completed 12 weeks of follow-up treatment. To evaluate the improvement in symptoms over time, we computed the item-wise difference between each subject’s symptom scores at baseline versus 12 weeks (ΔS). A positive value indicates an improvement in symptoms over the course of treatment. We then performed a principal component analysis (PCA) on the resulting matrix, first scaling all variables to have unit variance across N=30 subjects. The results of this PCA, computed on the ΔS scores, are denoted as “PCAΔ” throughout. Significance testing was performed by permutation testing: patient order was randomly shuffled for each symptom variable and the PCA was recomputed. This was repeated 5,000 times to establish the null distribution. Principal components (PCs) in the observed data which accounted for a higher proportion of variance than expected by chance (p<0.05) were used for further analysis. These PCs are denoted as “PCΔ”.

### Symptom PCA Stability Analyses

We assessed the stability of the symptom PCAΔ solution using a leave-one-out validation. First, we performed a PCA using the ΔS data from 29 (i.e., N-1) subjects. We then used the loadings matrix from this PCA and the symptom data from the left-out subject to predict their PC scores. We repeated this 30 times, each time leaving out a different subject. For each leave-one-out PCA, we compared the loadings to the full sample PCA via Pearson’s correlation across 25 symptom measure loadings. We also compared the predicted PC scores for all N=30 subjects to their observed (full sample PCA) scores.

To evaluate replicability of the symptom PCA solution, we also performed split-half validation using non-overlapping samples. First, we split the sample into two random halves, each with n=15 subjects. We then performed a PCA in each half independently, and compared the loadings between the two PCA solutions using a Pearson’s correlation. This process was repeated 1,000 times.

### Neural Data Preprocessing

Structural and functional neural images were preprocessed with the Quantitative Neuroimaging Environment & Toolbox (QuNex) framework (as described in (44)). First, images were converted from raw DICOM format to NIFTI volume-based files. Data were then preprocessed using the Human Connectome Project (HCP) minimal preprocessing pipelines (MPP)(45). Briefly, the MPP steps were as follows: the T1-weighted structural images for each subject were aligned with linear and non-linear warping in a single step to the standard Montreal Neurological Institute-152 (MNI-152) brain template, using the FMRIB Software Library (FSL) linear image registration tool (FLIRT) and non-linear image registration tool (FNIRT)(46). The same transformation was then applied to the T2-weighted structural image. Next, the FreeSurfer “recon-all” pipeline was used to produce individual anatomical segmentations in the cortex and subcortex (47). Cortical pial and white matter boundaries were generated as surfaces and used to define the ‘cortical ribbon’; subcortical gray matter voxels were segmented into anatomical structures. Together they represented the structural image in the Connectivity Informatics Technology Initiative (CIFTI) volume/surface ‘grayordinate’ space for each individual subject (45). Functional BOLD images were first motion-corrected by rigidly aligning to the middle frame of every run via FLIRT in the initial NIFTI volume space. Distortion correction was performed using a pair of reversed phase encoding direction spin-echo field maps. Next, a brain mask was applied to exclude non-brain signal. Cortical BOLD data were then mapped to the CIFTI gray matter matrix by sampling from the anatomically-defined gray matter cortical ribbon and then aligned to the HCP atlas using surface-based nonlinear deformation (45). Subcortical voxels were aligned to the MNI-152 atlas using whole-brain non-linear registration. The Freesurfer-defined subcortical segmentation was then applied to isolate the subcortical grayordinates of the CIFTI space.

After the HCP MPP, movement scrubbing was performed (48,49). All frames in the BOLD resting-state time-series with possible movement-induced artifactual fluctuations in intensity were flagged using two criteria: either the frame displacement (the sum of the displacement across all six rigid body movement correction parameters) exceeded 0.5 mm (assuming 50 mm cortical sphere radius) and/or the normalized root mean square (RMS) (calculated as the RMS of differences in intensity between the current and preceding frame, computed across all voxels and divided by the mean intensity) exceeded 1.6 times the median across scans. Every frame that met one or both of these criteria were discarded from further preprocessing and analyses. Subjects with more than 50% frames flagged would be entirely excluded, although no subjects in the present dataset were excluded due to this criterion (mean across the sample was 2.80% frames flagged, standard deviation 5.06). A high-pass filter (threshold 0.008 Hz) was then applied to the BOLD data to remove low frequency signals due to scanner drift. Next, QuNex was used to compute: 1) the average variation in BOLD signal within the ventricles, deep white matter, and across the whole gray matter (i.e. global signal), 2) movement parameters, as well as 3) their first derivatives to account for delayed effects. These variables, which likely do not reflect neural activity (44,50), were regressed out of the BOLD time-series as nuisance variables. Using global signal regression remains the field-wide gold standard for removing spatially persistent artifactual signal, although it is an actively developing field (see alternative arguments at (51,52); and emerging approaches at (53,54)).

### Data Quality Control

Only subjects with complete behavioral data (BPRS and MCCB measures at both baseline and Week 12 time points) were considered in the analyses. Additionally, neural structural T1w and functional BOLD images were subjected to visual quality control by a highly experienced researcher to check the fidelity of surface segmentation and registration of structural and functional data. For each structural image, the placement of the gray and white matter surfaces was assessed, and the registration of the BOLD image to the T1w was visually checked. The first functional scan for each subject was used. The temporal signal-to-noise ratio (TSNR) was also calculated for each functional BOLD run, and scans with low TSNR (here defined as being 2 standard deviations below the mean of all scans) were also excluded from further analysis. A total of 8 subjects were excluded (2 for failing neural data quality control and 6 for missing behavioral data). A total of 30 subjects were included in the analyses.

### Cortical and Subcortical Parcellations

We used the Cole-Anticevic Brain Network Parcellation (CAB-NP version 1.1.0)(55), which leveraged the Human Connectome Project’s Multi-Modal Parcellation (MMP1.0) to define functional networks and parcels across the cortex and subcortex. The cortical component of the parcellation comprises 180 bilateral parcels (total 360), consistent with the HCP MMP1.0, each assigned to one of 12 functional networks based on resting-state functional connectivity (FC). The subcortical component comprises 358 volumetric parcels, defined using resting-state FC with the cortical networks (55). The CAB-NP can be accessed at the Brain Analysis Library of Spatial maps and Atlases (BALSA) resource (https://balsa.wustl.edu/rrg5v) as well as the public release repository (https://github.com/ColeLab/ColeAnticevicNetPartition).

### Global Brain Connectivity (GBC)

For parcel-wise GBC maps we first computed the mean BOLD signal within each parcel for each participant and then computed the pairwise FC between all parcels. Following preprocessing and parcellation, the functional connectivity (FC) matrix was calculated for each participant by computing the Pearson’s correlation between every region in the brain with all other regions. This order of operations has been previously shown to result in stronger statistical values (compared to if GBC is computed prior to parcellation) due to increased within-parcel SNR of the BOLD data (44). A Fisher’s r-to-Z transform was then applied. Global brain connectivity (GBC) was calculated by computing every parcel’s mean FC strength with all other parcels (i.e. the mean, per row, across all columns of the FC matrix). GBC is a data-driven summary measure of connectedness that is unbiased with regards to the location of a possible alteration in connectivity and is therefore a principled way for reducing the number of neural features while assessing neural variation across the entire brain. Specifically, GBC is computed by:

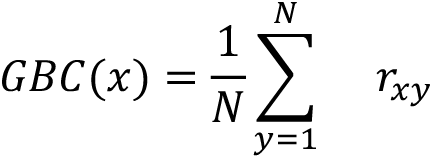

where *x* and *y* are parcels; *GBC*(*x*) denotes the GBC value at parcel *x*; *N* denotes the total number of parcels; and *r_xy_* denotes the correlation between the time-series of parcels *x* and *y*. We selected GBC because: i) the metric is agnostic regarding the location of dysconnectivity as it weights each area equally; ii) it yields an interpretable dimensionality-reduction of the full FC matrix; iii) unlike the full FC matrix or other abstracted measures, GBC produces a neural map, which can be related to other independent neural maps (e.g. gene expression maps, discussed below).

### Symptom-Neural Mapping

Subject-level behavioral scores for each PC, as well as total BPRS and MCCB scores, were mapped to individual dense GBC maps via mass univariate regression. Age, sex, and scanner were included in the regression model as covariates of no interest. The coefficients for behavior were then Z-scored for each map. The resulting map of Z values reflected the strength of the relationship between patients’ PC scores and the baseline GBC at every neural location, across all 30 patients. These regression coefficient maps are referred to as the βXGBC maps, where X is the behavioral score in the regression model (e.g. βPC1,Δ GBC for a regression of PC1Δ scores on to GBC).

### Symptom-to-Neural Mapping Stability Analyses

We assessed the stability of the symptom-to-neural regression in two ways: 1) We performed the mass univariate regression of PCΔ scores to GBC in N-1 subjects, repeating this 30 times and leaving out a different subject each time. 2) We performed the mass univariate regression to GBC of predicted PCΔ scores (from the leave-one-out PCA, see “Symptom PCA Stability Analyses” above). We did this across all 30 subjects (Figure 2) and repeated 30 times each time leaving one subject out (**Supplementary Fig. 6**). For each βPCGBC map, we correlated the β values with those from the corresponding observed (full sample) map across 718 parcels in the cortex and subcortex. These results assessed how sensitive the symptom-to-neural mapping is to the removal of one subject.

**Figure 1.**
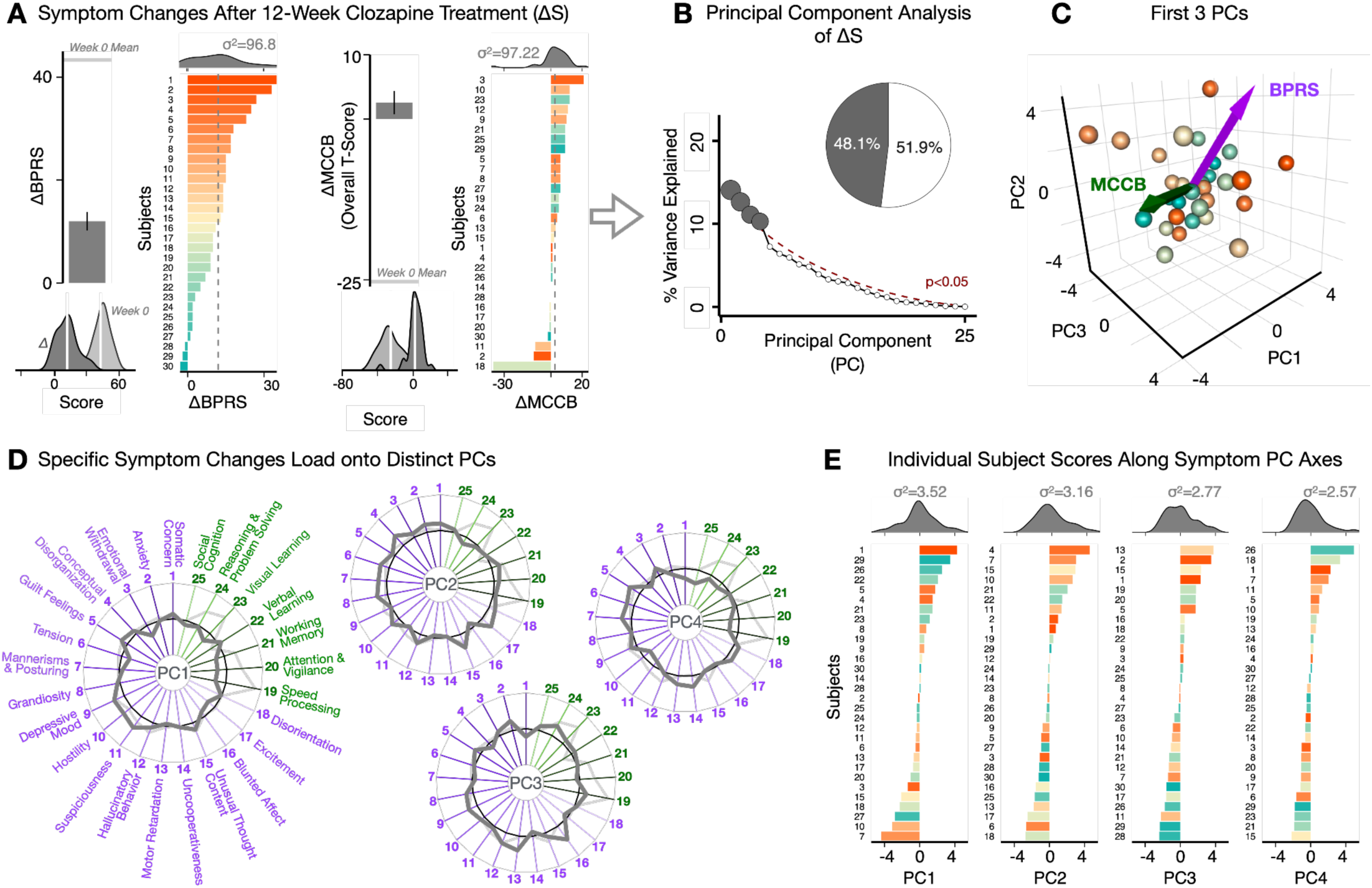
Dimensionality reduction of symptom change reveals distinct patterns of clozapine response. **A)** Differences between the total scores of core psychosis symptom measures (Brief Psychiatric Rating Scale, BPRS) and cognitive performance (MATRICS Consensus Cognitive Battery, MCCB), for N=30 subjects between baseline (week 0) and week 12 of the study. Note that the directionality of the MCCB scores was flipped such that a higher score corresponds with greater symptom severity (i.e. poorer cognitive performance), for consistency with the BPRS. Bar plots show group mean and standard error; ridgeline plots show distribution of scores across all subjects (white lines indicate group means). Ordered histograms show the improvement in scores over 12 weeks of clozapine treatment for individual subjects, (ΔBPRS and ΔMCCB respectively); each bar represents a single subject and a positive value indicates symptom improvement. Note that the color and label of each subject are preserved in the two histograms, showing that subjects with the most improvement in psychosis symptoms are not the same as those with the most improvement in cognitive functioning. The mean change in symptoms across all subjects is shown by the vertical gray dashed line. **B)** Screeplot shows the % variance explained by each of the principal components (PCs) from a PCA performed using the 12-week change in all 24 symptom measures across 30 patients (ΔS). The size of each point is proportional to the variance explained. The first four PCs (gray) survived permutation testing (p<0.05, 5,000 permutations). Together they capture 47.4% of all ΔS variance (inset). **C)** PCA solution shown in the coordinate space defined by the first three PCs. Colored arrows show the composite ΔBPRS and ΔMCCB vectors projected into the PC coordinate space. These vectors do not align with data-driven PC axes, highlighting that symptom improvement is not captured fully by changes in either BPRS or MCCB scores alone. Spheres denote individual subjects, colored as in panel A histograms. **D)** Loading profiles shown in dark gray for the 25 symptom measures on the 4 significant PCs. Each PC can be interpreted based on the pattern of loadings on individual symptom measures. **E)** Ordered histograms of individual subject PC scores. Bars represent individual subjects, colored as in panel A histograms. Distributions of PC scores are shown on the side.

**Figure 2.**
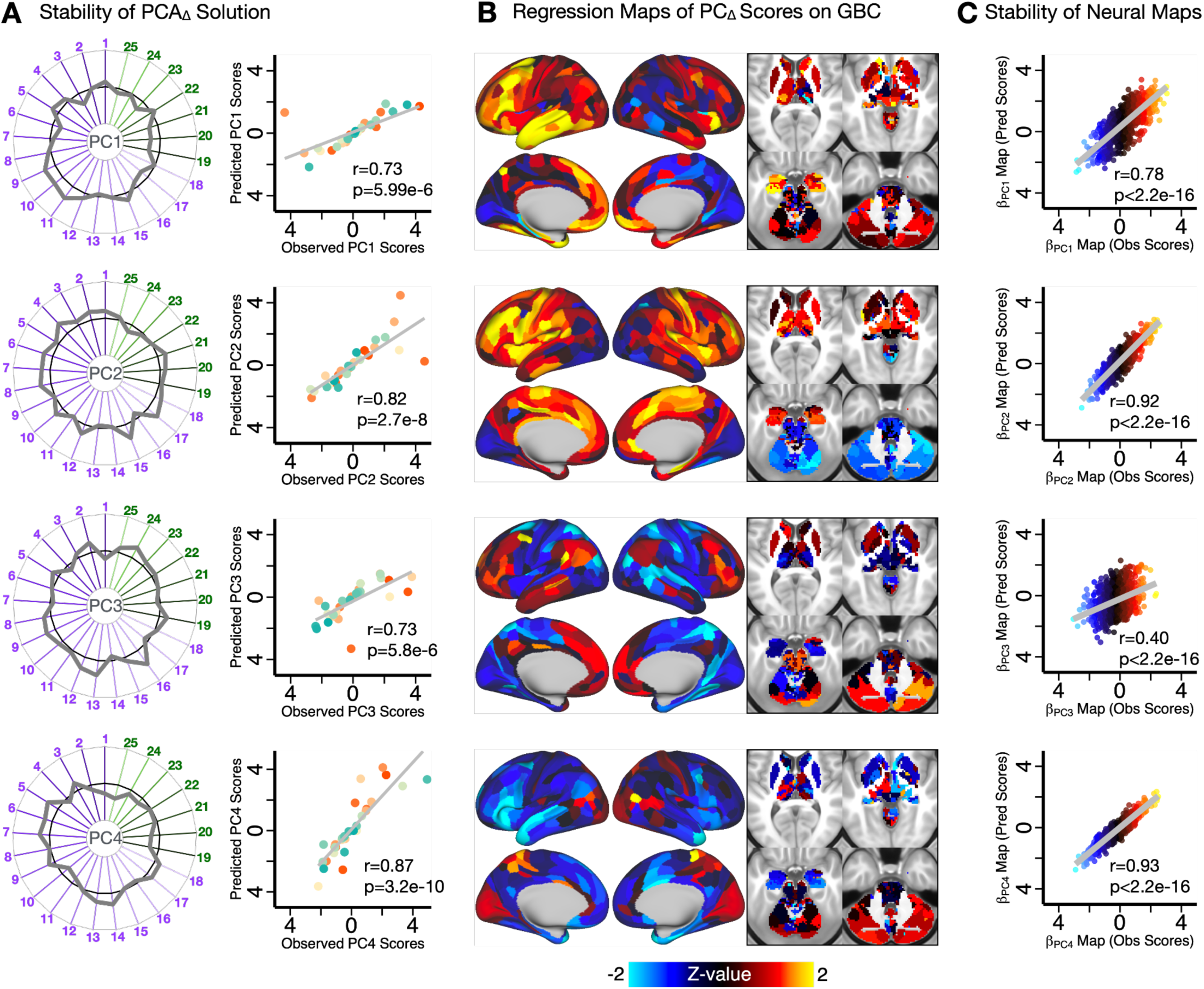
Symptom-neural mapping using dimensionality-reduced symptom scales reveal robust maps. **A)** Leave-one-out validation of the PCA_Δ_ symptom solution revealed that the PCA results are robust (see **Methods**). Radarplots show the loading profile for each PC. Scatterplots show the correlation between observed (full sample PCA solution) and predicted (from leave-one-out PCA) subject scores across N=30. **B)** β_PC,Δ_GBC from mass univariate regression of dimensionality-reduced PC scores onto parcelwise GBC across all patients. **C)** Mass univariate regression of leave-one-out predicted (Pred) PC scores on to GBC across N=30 produces β_PC_ maps that are highly significantly similar to the observed (Obs) β_PC,Δ_GBC maps.

Additionally, we assessed replicability of the symptom-neural mapping using the symptom scores from 1,000 runs of split-half PCA. For each run, we performed the mass univariate regression in both half-samples and then compared the two half-sample βPCGBC maps using Pearson’s correlation.

### Dimensionality Reduction of Neural Maps

We conducted a PCA on the parcellated neural baseline GBC maps across 30 subjects. This resulted in 29 neural PCs, denoted as PCGBC. Each map represents a principal component of neural variance, with the strength of the value in each parcel reflecting its loading on to that component. As with the behavioral PCA, significance of the neural PCA solution was assessed via permutation testing (5000 random within-subject shuffles of parcels).

### Relationship with Clozapine

For relationships with clozapine, the modal prescribed dosage of clozapine over 12 weeks was used. As described in the Study Design, adherence to clozapine treatment was established with monthly blood clozapine levels and by patient self report during all research study visits.

### Gene Expression Mapping

The gene expression maps were derived using Gene Expression Mapping Integrated with Neuro-Imaging for Discovery Of Therapeutics (GEMINI-DOT) version 1.20.0 (accessed June 15 2021 at https://bitbucket.org/murraylab/gdot/src/master/). The process has previously been described in (35). Briefly, the cortical and subcortical gene expression data were obtained from the publicly available Allen Human Brain Atlas (AHBA, RRID:SCR_007416), which quantified the expression levels of 20,737 genes obtained from six postmortem human brains using DNA microarray probes sampled from hundreds of neuroanatomical loci. Prior studies demonstrated the ability to map the expression level of each of these genes onto neuroimaging-compatible templates such as the CIFTI hybrid surface-volume format (34,35). In line with recently published methods (35), we mapped gene expression onto 180 symmetrized cortical parcels from the HCP MMP atlas (56) and 358 corresponding subcortical parcels from the CAB-NP (55). This yielded a map for each gene where the value in each parcel reflected the average expression level of that gene in the six brains in the AHBA dataset.

### Significance Testing of Gene Expression Maps

As done in our previous work, we first excluded any gene expression maps where the differential stability (DS) value was less than 0.1 (35,44). This included the HTR2B map (DS=0.06) and the DRD4 map (DS=0.06) To assess the significance of the correlation between gene expression maps and the target maps (i.e. βPC maps), we generated a null distribution of correlation values for each gene expression map and 100,000 surrogate maps which preserved the spatial autocorrelation characteristics of the target map. Surrogate maps were generated using the brainsmash software version 0.11.0 (https://github.com/murraylab/brainsmash) described in detail in (57). Using brainsmash, we first generated 100,000 surrogate maps for each target map, whose spatial autocorrelation was matched to the spatial autocorrelation of the target map. The surrogate maps were generated separately for the left and right cortical hemispheres and the subcortical volumes, then merged together to produce whole-brain surrogate maps. For each selected gene, we compute the correlation between the gene expression map and each of the 100,000 surrogate maps to build a distribution of 100,000 simulated r-values. We then used this distribution of simulated r-values to calculate the significance of the observed correlation between the gene expression map and the target map. All p-values were FDR-corrected across the selected genes. Finally, we selected genes whose observed r-value exceeded the distribution of 100,000 simulated r-values.

## RESULTS

### Data Reduction of Symptom Improvement Reveals Distinct Data-Driven Dimensions of Clozapine Response

We hypothesized that responses to clozapine would differ substantially between individual patients, i.e. that there would not be a uniform profile of symptom improvement over the 12 weeks of treatment. First, we characterized the change in symptoms between Week 0 and Week 12 (denoted by ΔS) in all 30 patients, using composite psychopathology and cognition symptom severity scores measured by the total BPRS and MCCB respectively (**Fig. 1A**). On average, patients improved by 12 points in total BPRS score (relative to the mean score of 43.33 at Week 0) and 16.2 points in total MCCB (relative to -243.9 at Week 0). Notably, patients with the greatest improvement in BPRS scores were not the same as those who improved in MCCB scores, as shown by the mismatched ordering between the histograms. Thus, we then attempted to compute the dominant dimensions of symptom improvement while agnostic to the BPRS and MCCB instrument constructs. These dimensions can be captured by the linear combinations of individual symptoms along which patients improved the most. Furthermore, we observed collinearity between individual ΔS measures (**Supplementary Fig. 1**), indicating that a dimensionality reduction of the symptom space may be sufficient to retain meaningful clinical variance.

We performed a principal component analysis (PCA) of the ΔS symptom data (PCAΔ), as this approach produces deterministic and orthogonal axes of maximal symptom change variation. This PCA captured four dominant modes that explained significantly more variance than would be expected by chance, totalling 48.1% of the overall behavioral variance (**Fig. 1B**). These four significant principal components (PCs) are a linear combination of the individual symptom items and are oblique to both the mean ΔBPRS and ΔMCCB axes, shown in **Fig. 1C**, indicating that the composite BPRS and MCCB scores do not optimally capture dimensions of symptom change over the 12 weeks of treatment. This is supported by further examining the individual loadings on each of the four PC axes (**Fig. 1D**), revealing that the dominant modes of clozapine-induced symptom change reflect symptoms related to paranoia (PC1Δ), positive and cognitive spectrum (PC2Δ), anxiety and negative symptoms (PC3Δ), and diffuse symptoms (PC4Δ). Furthermore, histograms of the individual subject scores along each of the PC axes show that individual patterns of symptom response to clozapine treatment is highly variable between patients, as hypothesized (**Fig. 1E**). Taken together, these results suggest that a data-driven dimensionality reduction solution can reveal individualized patterns of symptom improvement underlying rapid response to clozapine.

We next assessed the stability of the symptom PCAΔ solution. We opted to use leave-one-out validation given the relatively small number of subjects in the sample (see **Methods**), and show that all four PCAΔ symptom axes are robust to the removal of one subject (**Fig. 2A** and **Supplementary Fig. 6A**). However, to evaluate replicability of the solution, we also performed 1000 runs of split-half validation using non-overlapping samples (see **Methods**), The independent split-half solutions were moderately reproducible (**Supplementary Fig. 6C**). This is likely because the variance structure of each half-sample is determined by only a small number of subjects (n=15) which do not encapsulate the range of variance in the full sample. The robustness of the leave-one-out results suggest that the PCAΔ solution is stable and we expect that the split-half correlations will improve when sample sizes are increased.

### Residual Baseline Symptoms Remain after 12 Weeks of Clozapine Treatment

The PCAΔ revealed dimensions of maximal symptom change in response to clozapine. However, this analysis does not test whether the structure of dominant symptom dimensions at baseline would differ from those in the PCAΔ solution. We hypothesized some portion of the baseline symptom variance would not be reduced after 12 weeks of clozapine, and would therefore still be present in the Week 12 symptom geometry.

We conducted a PCA in both the baseline symptom data and the Week 12 data independently. At baseline, patients were on average moderate to severely symptomatic in both psychopathological symptoms and cognitive performance, as scored by the BPRS and MCCB (**Supplementary Fig. 2A**). This observation is expected as, in order to meet criteria for clozapine therapy, participants had to have responded poorly to two or more other antipsychotic drugs and show moderate to severe psychotic symptoms despite prior antipsychotic treatment. However, histograms of the individual-level scores reveal that the patients with the most severe BPRS symptoms are not the same as those with the most severe cognitive deficits. As with the ΔS difference scores, we computed a PCA of the SWeek0 baseline symptom data (PCAWeek0) in order to characterize the principal dimensions of symptom variation at baseline. This captured three significant axes, loading on to symptoms of cognitive impairment and disorganized thinking (PC1Week0), paranoia (PC2Week0), and negative symptoms (PC3Week0) respectively (**Supplementary Fig. 2**). Notably, patients with the highest baseline BPRS scores were also the ones who showed the most improvement over 12 weeks (r=0.65, p=9.6e-05, **Supplementary Fig. 2F**), possibly reflecting a regression to the mean; however, this relationship was markedly weaker for MCCB scores (r=0.22, p=0.23). Consistent with baseline results, patients with the lowest BPRS scores at Week 12 were also the ones who showed the most improvement over 12 weeks (r=0.48, p=0.0066, **Supplementary Fig. 3F** while the effect was moderate for MCCB scores (r=-0.27, p=0.15).

Importantly, we observed that the PCA of Week 12 symptoms (PCAWeek12, **Supplementary Fig. 3**) revealed a dominant PC1Week12 that is quantitatively similar to PC1Week0, both in its symptom loading profile (**Supplementary Fig. 4A**) and individual subject scores along this axis (**Supplementary Fig. 4B**). This suggests that the “cognitive impairment and disorganized thinking” symptom dimension improves relatively little under 12 weeks of clozapine treatment, as the symptom variance along this dimension remains present at Week 12. PC3Δ shows a similar, but weaker, effect with PC3Week0. However, the second component is notably different in the Baseline and Week 12 solutions, indicating that the symptom variance along this axis (which captures paranoia and positive symptoms at baseline) changes substantially between these two time points. In support of this, PC2Week0 is highly correlated with PC1Δ (**Supplementary Fig. 4C-D**).

### Relationship between Symptoms, Symptom Improvement, and Clozapine Dosage

We observed a trending negative relationship between modal clozapine dosage and the change in total BPRS score over 12 weeks (r=-0.36, p=0.078, **Supplementary Fig. 5A**), consistent with other reports (58). As this was not a fixed-dosage study, patients who did not show a reduction in symptoms at lower doses of clozapine were prescribed higher dosages. No significant relationships were found between modal clozapine and change in MCCB scores, or with baseline symptoms, residual Week 12 symptoms, or PCΔ scores (**Supplementary Fig. 5B-D**).

### Axes of Symptom Improvement Reveal Associations with Distinct and Specific Neural Systems

We next hypothesized that the dimensionality-reduced symptom change solution may improve the specificity of neural mapping as compared to pre-existing composite symptom scales. Specifically, we tested if the dimensionality-reduced axes of symptom improvement could identify novel and specific patterns of neural variation. Here we used parcel-wise global brain connectivity (GBC), a summary metric of functional connectivity, to capture neural variance as done in our previous publications. Parcellated GBC is a parsimonious measure reflecting how globally coupled an area is to the rest of the brain (59) that reduces the dimensionality of functional connectivity data in a neurobiologically grounded manner. Additionally, GBC is sensitive to alterations in cohorts with psychiatric symptoms, including in the psychosis spectrum (44,60–62) as well as in models of psychosis (63,64).

We find that each PCΔ dimension is associated with specific neural circuits (**Fig. 2B**). Higher PC1Δ (paranoia) scores are associated with elevated GBC in frontoparietal, language, and auditory networks, as well as ventral multimodal and default mode networks; PC2Δ (positive and cognitive spectrum) is associated with reductions in GBC dorsal attention network and elevations in somatomotor and auditory, as well as cingular-opercular regions; PC3Δ (anxiety and negative symptoms) shows reduced GBC across sensorimotor networks and reductions in dorsal attention, multimodal, and orbito-affective regions; while PC4Δ (diffuse symptoms) is associated with primarily reductions in GBC of language, default mode, and ventral multimodal regions. We also quantified the neural effects for ΔBPRS and ΔMCCB scores (**Supplementary Fig. 7**). While these maps, in particular the βΔBPRS map, showed strong effects and similarities with the βPC,Δ maps (**Supplementary Fig. 1****2**), these neural maps may reflect less specificity in symptom-associated circuits, as discussed below. **Supplementary Fig. 8** and **9** further show the neural effects of baseline and residual Week 12 symptom load respectively. Age, sex, and site were included as covariates in the symptom-neural regression model; the effects of these variables on GBC across subjects are shown in **Supplementary Fig. 10**.

We additionally examined the stability of the symptom-neural relationships: leave-one-out analysis shows that these maps are also highly robust (**Fig. 2C** and **Supplementary Fig. 6B/D**), although the βPC3,ΔGBC map appears to be only moderately stable compared to the other three maps. Importantly, dimensionality reduction of the neural GBC maps (see **Methods**) revealed three significant dominant components of connectivity variation across subjects that do not correlate with maps reflecting symptom change PCs (**Supplementary Fig. 1****1**), indicating that circuits with maximal neural connectivity variance at baseline do not necessarily track symptom response to clozapine. Thus, it is necessary to consider dimensions of symptom change in tandem with neural data, in order to identify neural systems that track clozapine response. The relationships between all neural maps are quantified in **Supplementary Fig. 1****2**. Taken together, these results suggest that different dimensions of symptom change over clozapine treatment are specifically associated with distinct neural systems in the brain. In comparison, the network effects for ΔBPRS and ΔMCCB scores (**Supplementary Fig. 7**), while also robust, are widespread across the brain. Thus, we next investigated whether the βPC,Δ neural maps are able to more specifically identify molecular targets associated with clozapine response and symptom improvement, compared to the maps of βΔBPRS and βΔMCCB.

### Informing Clozapine’s Therapeutic Targets via Gene Expression Mapping

Clozapine binds to a broad array of targets, including dopaminergic, muscarinic, adrenergic, histaminergic, and serotonergic receptor subtypes (31–33,65). These receptors, as well as other proteins involved in their mechanisms of action, are differentially expressed across the brain. However, it is unclear which of these targets are responsible for clozapine’s relatively superior therapeutic effects when compared to other antipsychotic medications; it is also possible that clozapine’s efficacy is due to a combination of binding targets, with different mechanisms responsible for different aspects of the observed clinical response (65). Importantly, some of these targets may also be responsible for clozapine’s side effects (65–67). Thus, identifying the mechanisms which underlie clozapine’s therapeutic effects may serve to inform the development of novel drugs with the same or better efficacy and potentially fewer side effects.

Here we leverage recent advances in human neural transcriptomic mapping (35) to inform the pharmacology underlying clozapine’s therapeutic effects. We related the spatial expression topographies of genes encoding clozapine’s receptor targets with the neural βΔ maps of clozapine-induced symptom change. First, we hypothesized that the βΔMCCB map would not show a strong association with receptor gene expression maps, reflecting the mild clinical effect and consequent attenuated neural effect as seen in **Supplementary Fig. 7**. Second, we hypothesized that the βΔBPRS map would show strong but non-specific associations with a broad range of receptors; this is because the clinical response measured by the ΔBPRS score (and the associated neural circuitry) captured broad changes in symptoms that are not selective to any specific dimension. Lastly, we hypothesized that the βPC,Δ maps, which reflect specific and orthogonal dimensions of clozapine-induced symptom change, would each be selectively associated with a subset of the expression maps of genes coding for specific receptors in clozapine’s binding profile. This would thus inform the targets underlying particular dimensions of therapeutic effects. Specifically, we reasoned that βPC,Δ maps should be highly spatially similar to the gene expression maps of receptors implicated in the mechanisms underlying the associated symptom changes; in contrast, βPC,Δ maps should not be significantly similar to the expression of receptors which are not directly involved in producing the observed symptom changes. For this analysis we considered the expression profiles of 23 genes encoding receptors implicated in clozapine’s neuropharmacology, including dopaminergic (DRD1, DRD2, DRD3, DRD5), serotonergic (HTR2A, HTR2C), adrenergic (ADRA1A, ADRA1B, ADRA1D, ADRA2A, ADRA2B ADRA2C), cholinergic (CHRM1, CHRM2, CHRM3, CHRM4, CHMR5), and histaminergic (HRH1, HRH2, HRH3, HRH4) receptor subunits.

As hypothesized, we observed that the βΔMCCB map did not significantly correlate with any of the gene expression maps of interest (**Fig. 3** and **Supplementary Table 2**). In contrast, the βΔBPRS map showed strong but broad significant relationships with most of the genes of interest, suggesting that while the BPRS may be well suited to capturing general changes in response to treatment, it may not be sufficiently sensitive to detect heterogeneous neurobiological responses to clozapine. The improvement in BPRS scores may be associated with a broad range of receptors rather than selective targets (see **Discussion**). Thus, we tested if the neurobehavioral PCΔ axes show specificity of neural gene expression, therefore capturing individual variation in clozapine response that can be related back to neural features.

**Figure 3.**
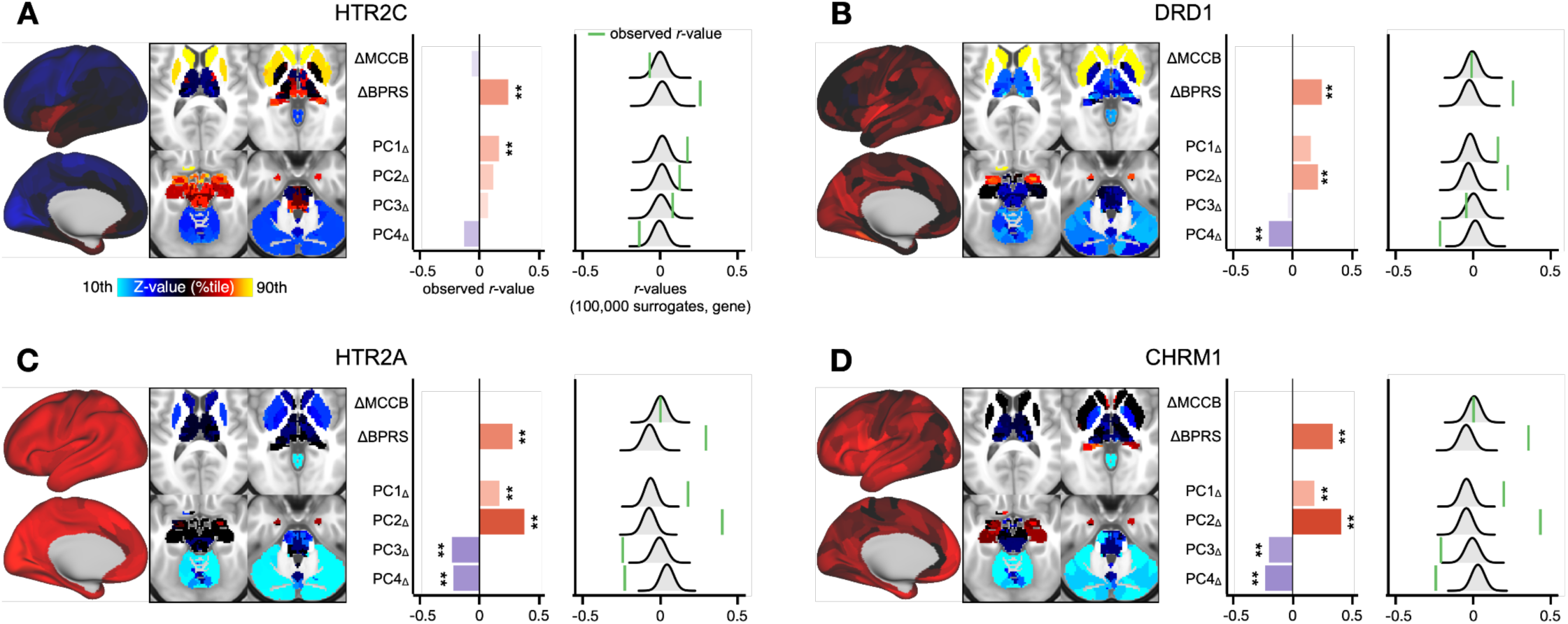
Informing clozapine’s therapeutic effects through gene expression mapping. Gene expression maps are derived from transcriptomic data from the AHBA (34,35). **A)** The map of HTR2C (which encodes the serotonin 5HT_2C_ receptor) expression is significantly correlated with the map of PC1_Δ_ symptom change (r=0.174, p=0), as well as the map of ΔBPRS symptoms (r=0.256, p=0). For each β_Δ_ map, significance was assessed by computing a null distribution of r-values between the gene expression map and 100,000 surrogate maps preserving the spatial autocorrelation of the original β_Δ_ map, as shown in the distributions. Type-I error protection was performed via a Bonferroni correction across all Gene vs. β_Δ_ map correlations. **B)** DRD1, which encodes the dopamine D_1_ receptor, is significantly correlated with the PC2_Δ_ (r=0.225, p=0) and the ΔBPRS map (r=0.259, p=0) and anti-correlated with PC4_Δ_ (r=-0.212, p=0). **C)** HTR2A, which encodes the serotonin 5HT_2A_ receptor, is significantly correlated with all PC_Δ_ maps, the Mean map, and the ΔBPRS map. **D)** CHRM1, which encodes the muscarinic acetylcholine receptor M_1_, is significantly correlated all except the ΔMCCB map.

We observed that each βPC,Δ map was significantly spatially correlated with a number of receptor gene expression maps (**Supplementary Table 2**), revealing higher sensitivity to variation in gene expression patterns relative to the ΔMCCB and ΔBPRS measures. Several genes were selectively correlated with specific βPC,Δ maps. For example, the map of HTR2C (which encodes the serotonin 5HT2C receptor) expression is significantly correlated with the βPC1,Δ map, but not any of the other βPC,Δ maps (**Fig. 3A**). While this analysis cannot establish causality, the highly significant relationship between the neural circuits associated with PC1Δ and the expression profile of 5HT2C receptors suggests that this receptor may be involved in the mechanism of action underlying changes in the PC1Δ “paranoia” symptom dimension (see **Discussion**). In several cases, the βPC,Δ maps were anti-correlated with select gene expression maps. For example, the DRD1 expression map, which encodes the dopamine D1 receptor, is significantly positively associated with the βPC2,Δ map and significantly negatively associated with the βPC4,Δ map, but not βPC1,Δ or βPC3,Δ (**Fig. 3B**). These results point to D1 receptors as a possible primary target involved in PC2Δ “positive and cognitive spectrum” and PC4Δ “diffuse” symptom changes but with potentially opposing effects on these two symptom dimensions.

Notably, some genes were also broadly associated with all βPC,Δ symptom maps, e.g. HTR2A (which encodes the 5HT2A receptor, **Fig. 3C**) and CHRM1 (which encodes the muscarinic acetylcholine receptor M1, **Fig. 3D**). These receptors may be broadly involved in changes in several symptom domains.

## DISCUSSION

Clozapine is currently the only antipsychotic with demonstrated efficacy for TRs (7–9). However, due to the associated risk of agranulocytosis and lack of patient selection strategies to determine likelihood of maximal early response (14), clozapine remains under-prescribed in clinical practice. Only about 3.5% of patients with TRS in the US receive a trial of clozapine (13,68), although it has been estimated that up to 25-30% of all patients would benefit from clozapine (69). Predictive clinical or biological indicators that can inform a given patient’s response to clozapine could greatly increase its early use in patients most likely to benefit from this intervention. Moreover, understanding the neuropharmacology of clozapine in relation to its heterogeneous therapeutic effects will also inform new symptom-relevant neural targets for which novel therapies can be optimized.

Here we evaluated whether patterns of clinical clozapine response in TRs can be quantitatively related to neural functional connectivity alterations in patients who were imaged prior to or right at the onset of clozapine treatment. First, we showed that data-reduction of the item-level clinical scales revealed distinct axes of clozapine response patterns. Critically, these data-driven symptom patterns of clozapine response were related to baseline neural maps. In that sense, the variation across patients in their GBC patterns can be considered as a possible neural failure mode that can be mapped to the data-driven clozapine clinical response pattern and used to stratify patients who are most likely to respond to clozapine treatment (38). Consistent with this notion, we found that relating clinical dimensions of symptom change to individual GBC maps revealed stable patterns (despite the modest sample size) that were distinct from simply correlating the mean improvement on any one clinical scale. Put differently, we identified dimensions of clinical response mapped onto neuroimaging features most related to individual patterns of symptom change. We further studied possible receptor targets that map onto neuro-behavioral clozapine response patterns via neural gene expression derived from the Allen Human Brain Atlas (35). Here we focused on receptor targets implicated in clozapine’s pharmacology and show that different data-driven neuro-behavioral clozapine response maps are quantitatively related to distinct receptor targets. Collectively, these results suggest that mapping individual variation of pharmacological clinical and neural patterns reveal potential gene-expression informed targets for maximal individualized clozapine response optimization.

### Dominant Dimensions of Individual Clinical Clozapine Response Map to Neural Features

Our dimensionality reduction analysis revealed four significant dimensions of symptom change over 12 weeks of clozapine treatment, corresponding to axes along which patients showed the greatest improvement. These dimensions highlight the heterogeneity of individual response to clozapine, showing graded and distinct patterns of change in paranoia, positive symptoms and cognitive performance, anxiety and negative symptoms, and diffuse symptoms even within a sample of 30 TRS patients. Importantly, the dimensionality reduction solution reveals that the dominant axes of symptom change in this cohort do not map exactly onto improvements in composite BPRS (and/or MCCB) scores. Rather, patients show distinct patterns of symptom change, and even individuals who did not improve in their total BPRS score showed varying degrees of improvement in specific symptoms. This suggests that the total BPRS score, which is often used to define clinical treatment response in TRS (70), may not be specific enough to capture nuanced behavioral improvements in response to clozapine. This is further supported in our results by the widespread neural systems and receptor systems associated with an improvement in total BPRS score (**Supplementary Fig. 7**, **Supplementary Table 2**). Here we present evidence for dissecting the mechanistic and therapeutic underpinnings of specific axes of behavioral treatment response by first parsing the symptom heterogeneity observed in TRS.

Interestingly, while cognitive symptoms showed some improvement in this cohort as shown by the loadings of cognitive items on several of the PCΔs (**Fig. 1**), these symptoms represented the principal axis of variance in both the PCWeek0 and PCWeek12 solutions (**Supplementary Fig. 2**-**3**). This suggests that a large proportion of the variance attributed to cognitive deficits remain unaffected over the course of clozapine treatment (71), highlighting the need to develop novel treatments for targeting cognitive symptoms in TRS.

### Using Gene Expression Maps to Identify Mechanisms for Targeting Selective Symptoms

Clozapine uptake is widespread across the brain and has a broad range of receptor targets, including dopaminergic, muscarinic, adrenergic, histamine, and serotonergic receptor subtypes (31–33). To study whether distinct neural targets are associated with different dimensions of clinical clozapine response, we quantified each of the symptom-relevant neural functional connectivity maps with the expression profiles of genes encoding various receptors of interest. We observed that βPC,Δ maps are associated with the spatial topographies of distinct receptor subtypes, suggesting that these differential patterns of targets may underlie the heterogeneous clinical effects of clozapine. Furthermore, while some receptor subtypes were selectively associated with specific symptom-relevant neural maps, others were more widely associated. Notably, compared to the βPC,Δ maps, the βΔBPRS map is correlated with many of the gene expression maps of interest (**Supplementary Table 2**). This suggests that the improvement in overall BPRS scores in this sample of TRS patients may be associated with a broad range of receptor targets. This may be because the BPRS battery is designed to capture broad symptoms of canonical psychopathology, which would likely in turn be attributable to many receptor targets and mechanisms of action. It is possible that while the BPRS composite is useful as a measure of overall clinical efficacy, it is less specific as an indicator of treatment response to clozapine, given the distinct dimensions of symptom change and the heterogeneity observed across individual responses to clozapine. Taken together, our results provide evidence for different neural targets that may underlie different aspects of clozapine response, and may serve as an opportunity to inform the development of optimized treatment for specific and targeted symptoms in schizophrenia.

Interestingly, the DRD2 gene expression map is not significantly correlated with any of the βPC,Δ, βΔBPRS, or βΔMCCB maps (**Supplementary Table 2**), possibly indicating that D2 receptors are not primarily responsible for the improvement in psychosis symptoms after clozapine treatment in this sample. This is in line with the lack of symptom improvement observed in these TRS patients after treatment with other antipsychotic medications, which act primarily through D2 receptor agonism (72). Additionally of note are the broad significant associations between symptom improvement maps and the expression of the muscarinic acetylcholine receptor M1 gene, CHRM1. Consistent with this observation, muscarinic receptor agonists such as xanomeline (which preferentially targets M1 and M4 receptors (73–75)) have been successfully shown to produce robust antipsychotic effects across symptom domains in preclinical animal models (76–78) and more recently in clinical trials (74,79).

### Considerations and Future Directions

This study serves as a small-scale proof-of-concept, which lays the groundwork for using state-of-the-art neuroimaging analytics and independent gene expression to inform the development of markers for early treatment decisions in TRS. However, there are several constraints on the current results that require further study and development. Firstly, while the results of the symptom PCA and neural mapping were stable in our sample of N=30 TRS patients, these effects must be shown to replicate in a larger independent sample. Secondly, as this was not a fixed-dosage study, the individual responses to clozapine may have been affected by differences in drug dosage, as well as differences in acclimating to dosage changes. To mitigate this potential confound, we chose to use the modal dosage, which for most patients was a steady amount; nevertheless, the impact of drug dosage can only be eliminated in a fixed-dose study with constant monitoring of clozapine levels. Unfortunately, a fixed-dose study of clozapine would be difficult to conduct given the need to start with very small doses to minimize the risk of significant adverse effects. In addition, there is substantial variability in the doses tolerated by patients, given the prominent side effect profile. Furthermore, our results point to the individual variation in both clinical and neural features of clozapine response and how this heterogeneity may be related to specific receptor pathways. Follow-up studies will be needed to study the heterogeneity of receptor response and enrich the information around neural features.

## Conclusions

We present evidence that resting state functional connectivity may serve as a prognostic biomarker of clozapine response, defined using empirically-derived neuro-behavioral measures, for patients with TRS. We show that distinct patterns of clozapine symptom response can be mapped to neuroimaging features, revealing symptom-relevant neural targets of clinical efficacy. Lastly, we quantified response-relevant neural maps against the expression profiles of receptor genes implicated in clozapine’s pharmacology, demonstrating that distinct dimensions of clozapine response can be associated with distinct receptor targets. We present a framework for identifying neuro-behavioral targets linked to pharmacological efficacy that can be further developed to inform optimal early treatment decisions in schizophrenia.

## Data Availability

Due to the clinical nature of the data, data will be shared only through request following a review of the project proposal. Depending on the proposal, a formal data sharing agreement and appropriate data safety measures may be required.

## Acknowledgments

This manuscript was funded by NIH Grants MH109508 (to AKM), 5P50AA012870 (to AA), 5R01MH108590 (to AA), 5R01MH112189 (to AA), 5R01MH115000 (to AA), 5R01MH116038 (to AA), 5R01MH112746 (to AA), 5U01MH121766 (to AA), 5R24MH122820 (to AA), and 5U01MH124639 (to AA).

## SUPPLEMENT

**Supplementary Figure 1.**
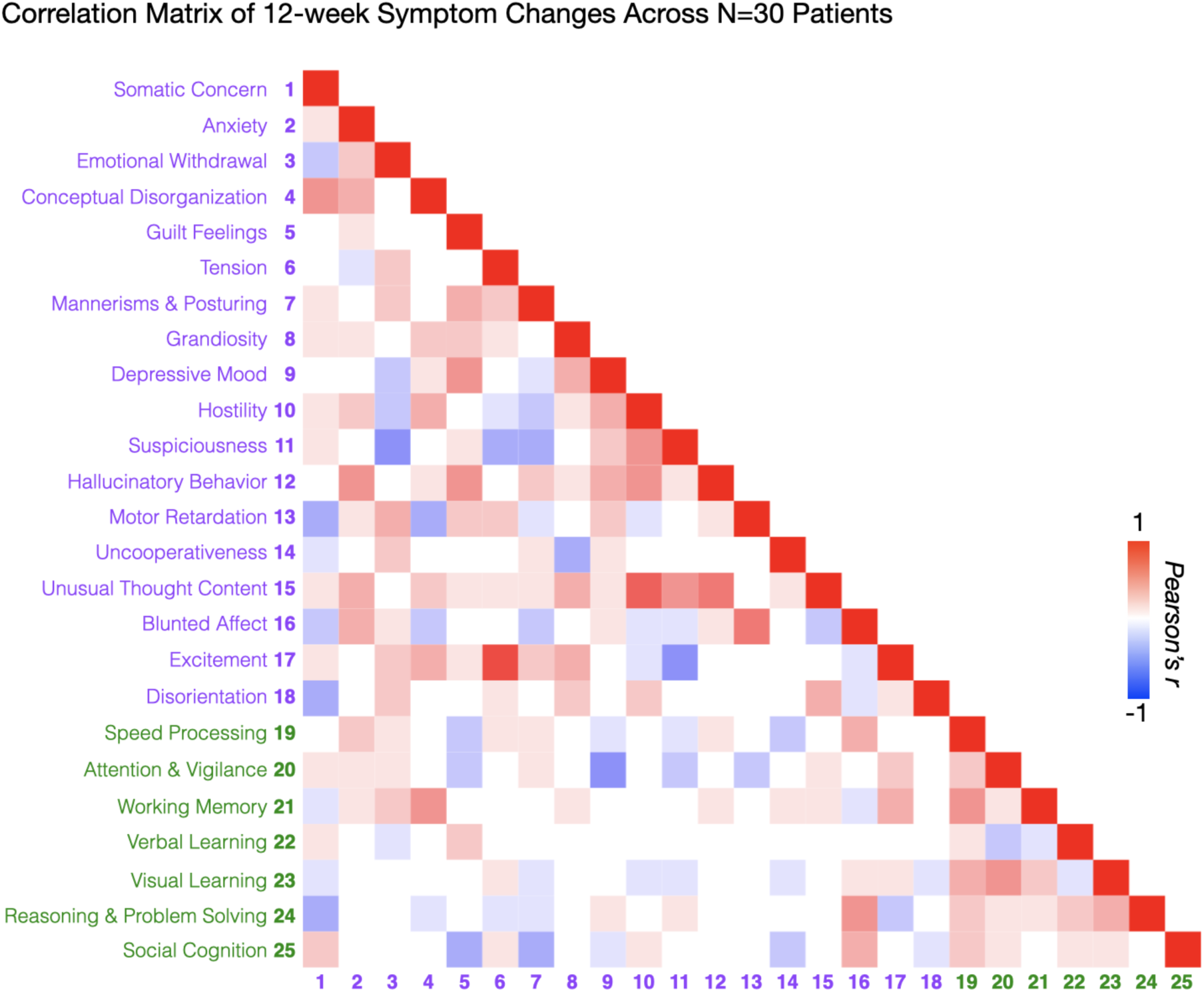
Correlation matrix of changes in individual symptom measures over 12 weeks of clozapine treatment.

**Supplementary Figure 2.**
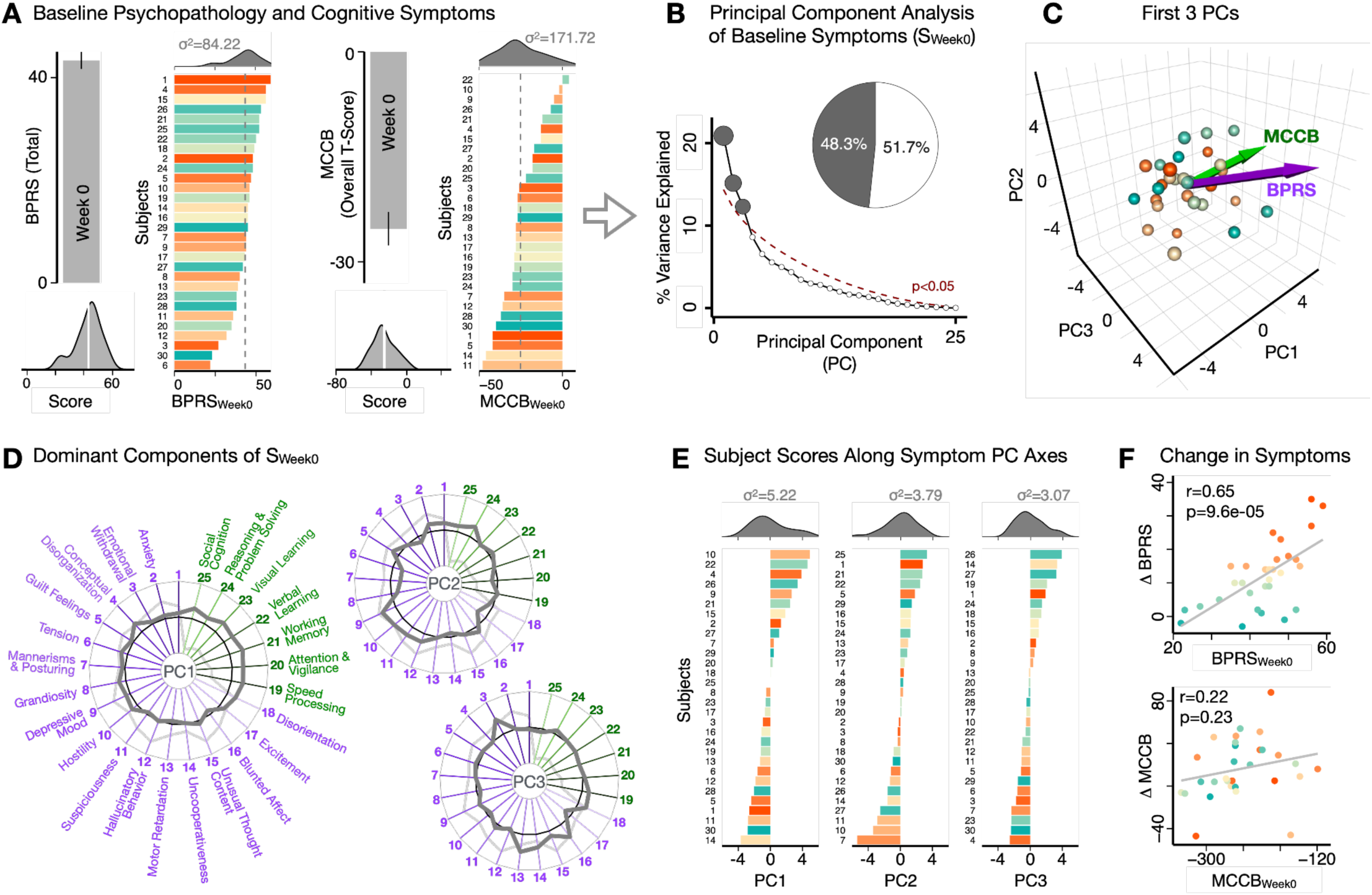
Dimensionality reduction of baseline symptoms reveals dominant modes of symptom variation in N=30 antipsychotic-resistant SCZ. **A)** Composite total core psychosis symptom measures (Brief Psychiatric Rating Scale, BPRS) and cognitive performance (MATRICS Consensus Cognitive Battery, MCCB), for N=30 subjects at baseline (Week 0) of the study. Note that the directionality of the MCCB scores was flipped such that a higher score corresponds with greater symptom severity (i.e. poorer cognitive performance), for consistency with the BPRS. Bar plots show group mean and standard error; ridgeline plots show distribution of scores across all subjects (white lines indicate group means). Ordered histograms show the total baseline BPRS_Week0_ and MCCB_Week0_ scores for individual subjects. Note that the color of each subject is preserved in the two histograms; subjects with the most severe psychosis symptoms are not the same as those with the most severe cognitive deficits. The mean total score across all subjects is shown by the vertical gray dashed line. **B)** Screeplot shows the % variance explained by each of the principal components (PCs) from a PCA performed using baseline symptoms in all 25 symptom measures across 30 patients (S_Week0_). The size of each point is proportional to the variance explained. The first three PCs (gray) survived permutation testing (p<0.05, 5,000 permutations). Together they capture 48.3% of all S_Week0_ variance (inset). **C)** PCA solution shown in the coordinate space defined by the first three PCs. Colored arrows show the composite BPRS_Week0_ and MCCB_Week0_ vectors projected into the PC coordinate space. Spheres denote individual subjects, colored as in panel A histograms. **D)** Loading profiles shown in dark gray for the 25 symptom measures on the 3 significant PCs. Each PC can be interpreted based on the pattern of loadings on individual symptom measures. **E)** Ordered histograms of individual subject PC scores. Bars represent individual subjects, colored as in panel A histograms. Distributions of PC scores are shown on the side. **F)** There is a significant correlation across subjects between the total BPRS_Week0_ score and the improvement in BPRS over 12 weeks of clozapine (r=0.65, p=0.6e-05), indicating that patients with the most severe BPRS symptoms at baseline also showed the greatest improvement. This relationship does not hold for the MCCB cognitive performance scores (r=0.22, p=0.23).

**Supplementary Figure 3.**
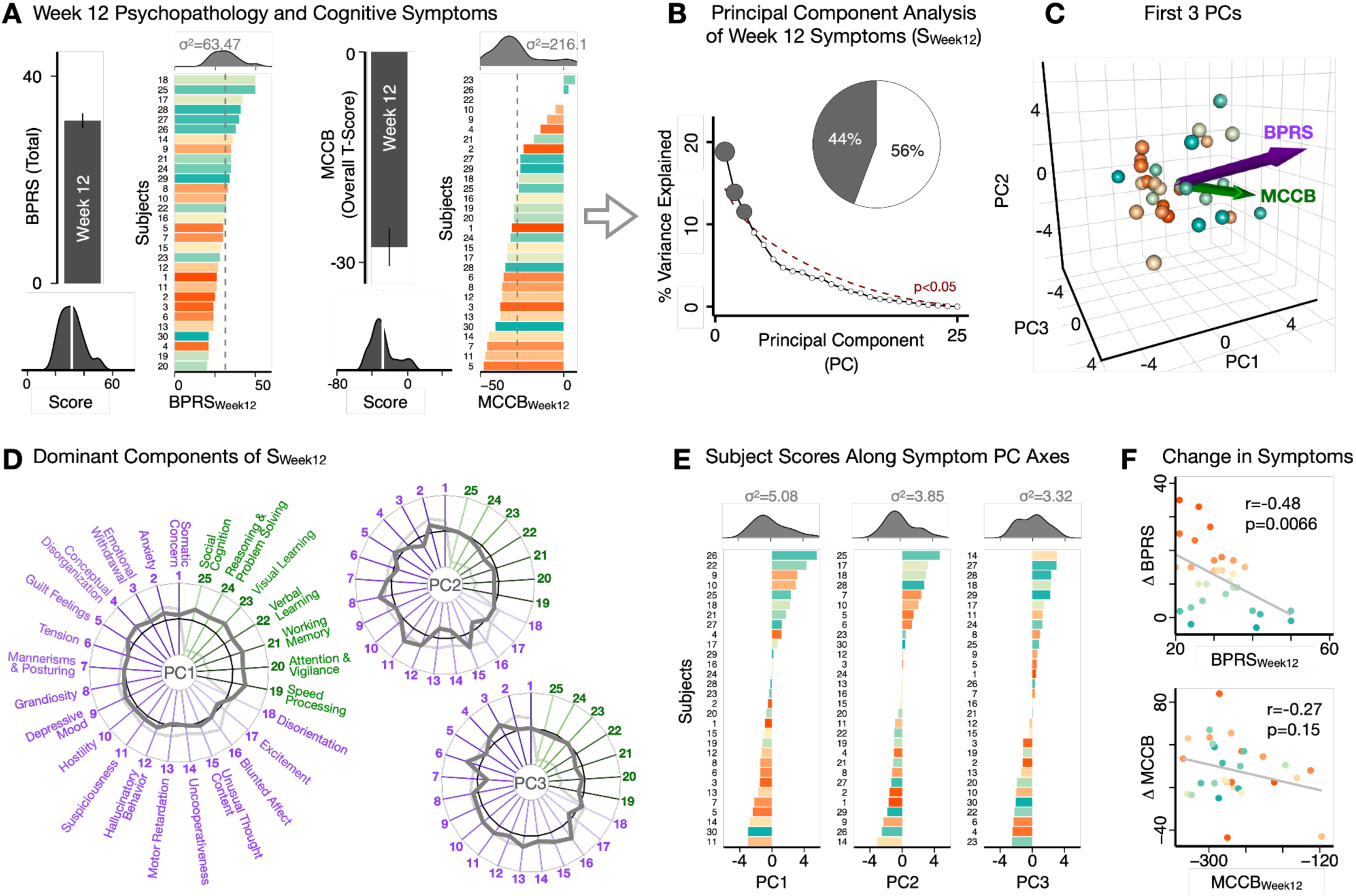
Dimensionality reduction of symptoms at Week 12 of clozapine treatment, revealing dominant modes of symptom load in N=30 SCZ. **A)** Composite total core psychosis symptom measures (Brief Psychiatric Rating Scale, BPRS) and cognitive performance (MATRICS Consensus Cognitive Battery, MCCB), for N=30 subjects at Week 12 of the study. Note that the directionality of the MCCB scores was flipped such that a higher score corresponds with greater symptom severity (i.e. poorer cognitive performance), for consistency with the BPRS. Bar plots show group mean and standard error; ridgeline plots show distribution of scores across all subjects (white lines indicate group means). Ordered histograms show the total BPRS_Week12_ and MCCB_Week12_ scores for individual subjects. Note that the color of each subject is preserved in the two histograms; subjects with the most severe residual psychosis symptoms are not the same as those with the most severe cognitive deficits. The mean total score across all subjects is shown by the vertical gray dashed line. **B)** Screeplot shows the % variance explained by each of the principal components (PCs) from a PCA performed using baseline symptoms in all 25 symptom measures across 30 patients (S_Week12_). The size of each point is proportional to the variance explained. The first three PCs (gray) survived permutation testing (p<0.05, 5,000 permutations). Together they capture 44% of all S_Week12_ variance (inset). **C)** PCA solution shown in the coordinate space defined by the first three PCs. Colored arrows show the composite BPRS_Week12_ and MCCB_Week12_ vectors projected into the PC coordinate space. Spheres denote individual subjects, colored as in panel A histograms. **D)** Loading profiles shown in dark gray for the 25 symptom measures on the 3 significant PCs. Each PC can be interpreted based on the pattern of loadings on individual symptom measures. **E)** Ordered histograms of individual subject PC scores. Bars represent individual subjects, colored as in **Fig. 1A** histograms. Distributions of PC scores are shown on the side. **F)** There is a significant negative correlation across subjects between the BPRS_Week12_ score and the improvement in BPRS over 12 weeks of clozapine (r=-0.48, p=0.0066), indicating that patients with the most severe residual BPRS symptoms at baseline also showed the least improvement. This relationship is attenuated for the MCCB cognitive performance scores (r=-0.27, p=0.15).

**Supplementary Figure 4.**
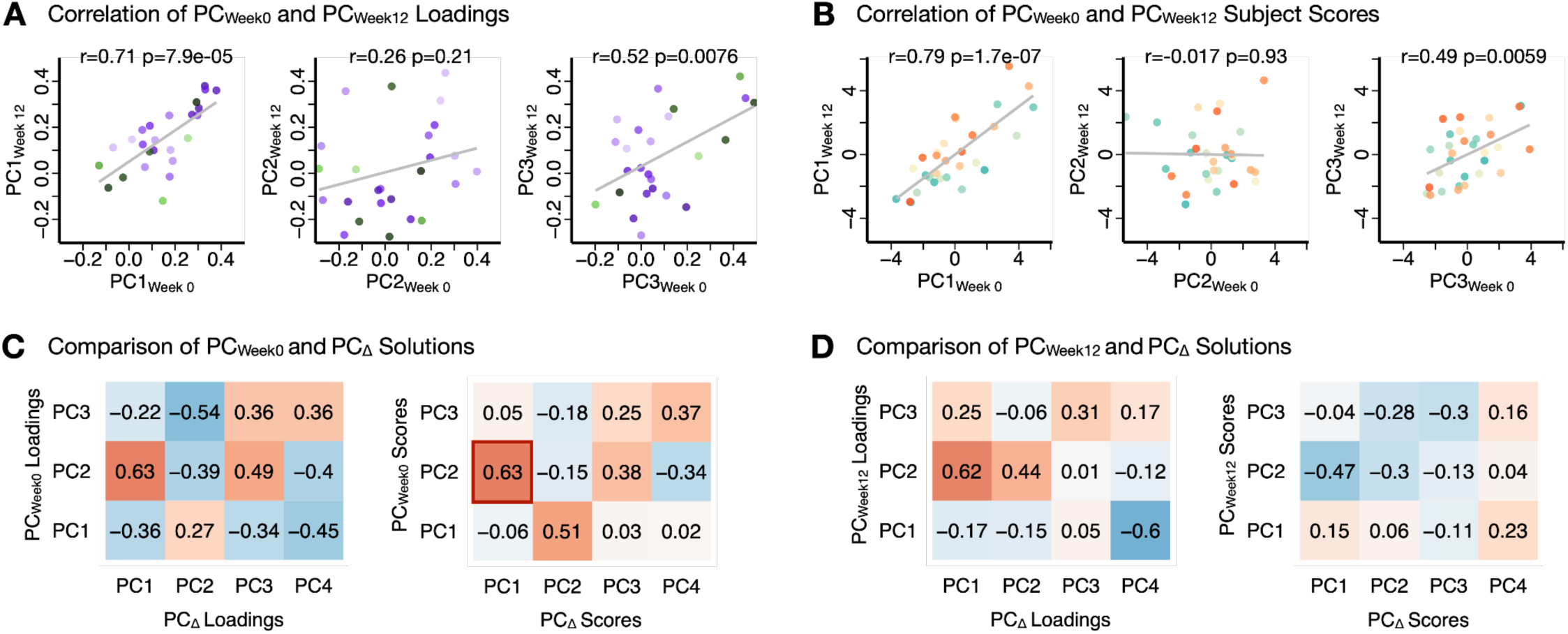
Comparison of the PC_Week0_, PC_Week12_, and PC_Δ_ solutions. **A)** Correlation of the loadings of 25 symptom measures between the PC_Week0_ and PC_Week12_ solutions. Each point corresponds to a symptom measure from the BPRS or MCCB. The loadings for PC1 are highly similar between baseline and Week 12 solutions (r=0.71, p=7.9e-05). **B)** Correlations of the subject scores between the PC_Week0_ and PC_Week12_ solutions. Each point corresponds to a subject. Again, PC1 is highly similar between the two PCAs, suggesting that this dimension has not significantly changed over the 12 weeks of clozapine in this sample. **C)** Correlations (r-values) between the loadings (left) and subject scores (right) of the PC_Week0_ and PC_Δ_ solutions. PC_Δ_ resembles highly the PC2_Week0_ solution both in symptom loadings and subject scores, suggesting that this dimension changes most during the 12 weeks of clozapine treatment in this sample. **D)** Correlations (r-values) between the loadings (left) and subject scores (right) of the PC_Week12_ and PC_Δ_ solutions.

**Supplementary Figure 5.**
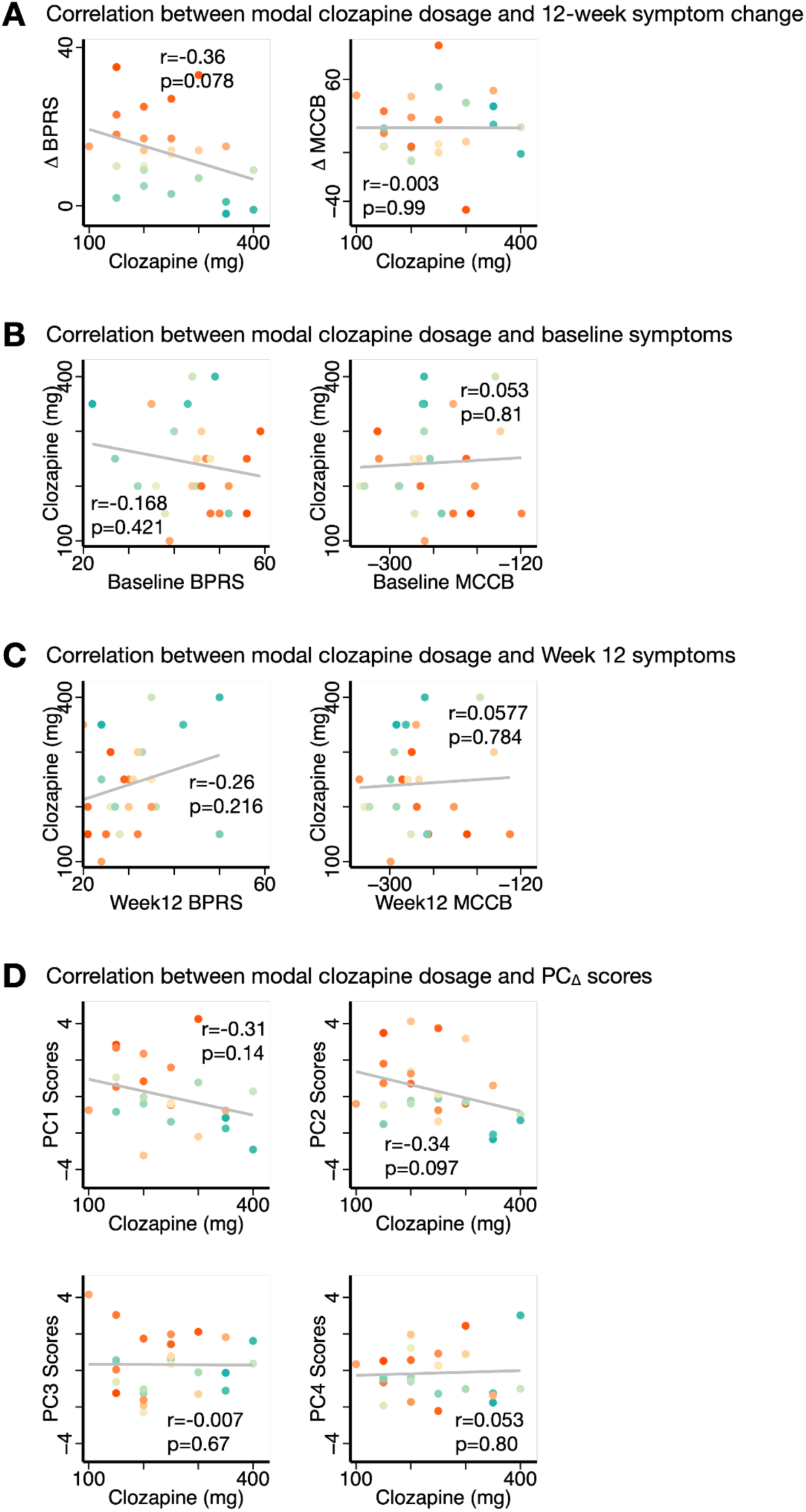
Relationships between behavior and clozapine dosage. **A)** Correlations between the modal clozapine dosage (mg) and change in BPRS and MCCB scores over 12 weeks of treatment across patients. As this was not a fixed-dosage study, the trending negative relationship between ΔBPRS and clozapine dosage is likely because patients who showed no improvement in BPRS to lower dosages were prescribed a higher amount of clozapine. **B)** Correlations between the modal clozapine dosage (mg) and baseline symptoms. **C)** Correlations between the modal clozapine dosage (mg) and residual symptoms at Week 12. **D)** Correlations between the modal clozapine dosage (mg) and PC_Δ_ scores.

**Supplementary Figure 6.**
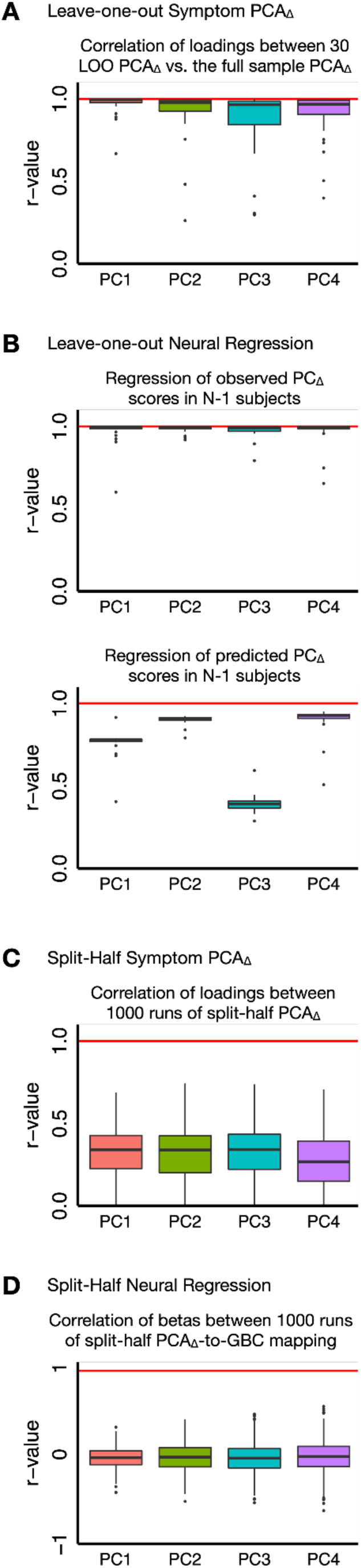
Assessing the stability of the PCA_Δ_ solution and symptom-neural mapping. **A)** Leave-one-out validation of the PCA_Δ_ solution. A PCA was conducted in N-1 subjects and the loadings were used to predict the PC scores of the left-out subject. The process was iterated over all 30 subjects. The correlation between loadings from all 30 PCAs and the full-sample PCA are shown (see **Methods**). **B)** Leave-one-out mapping of symptom scores to neural GBC. (Left) PC scores from N-1 subjects in the full-sample PCA solution were regressed onto GBC maps. Correlation values across parcels between the resulting β_PC,Δ_GBC maps and the full-sample β_PC,Δ_GBC maps are shown in the boxplot. (Right) Predicted PC scores for N-1 subjects, from the N-1 PCAs, were regressed on to GBC maps. Correlation values across parcels between the resulting β_PC,Δ_GBC maps and the full-sample β_PC,Δ_GBC maps are shown in the boxplot. **C)** Split-half validation of the PCA_Δ_ solution. For each run, the sample was split randomly into 2 even halves, each n=15, and a PCA was conducted in each half. This was repeated 1,000 times. The correlations between the loadings of the two PCAs in each split-half run are shown in the boxplot. **D)** Split-half validation of the symptom-neural mapping. For each of the 1000 runs of split-half PCA, the PC scores were regressed onto the respective GBC maps in each half-sample, producing two sets of β_PC,Δ_GBC maps. The correlations between the two split-half β_PC,Δ_GBC maps are shown in the boxplot. Across all boxplots, outliers are defined as data points more than (1.5 times the interquartile range) outside the upper or lower quartiles.

**Supplementary Figure 7.**
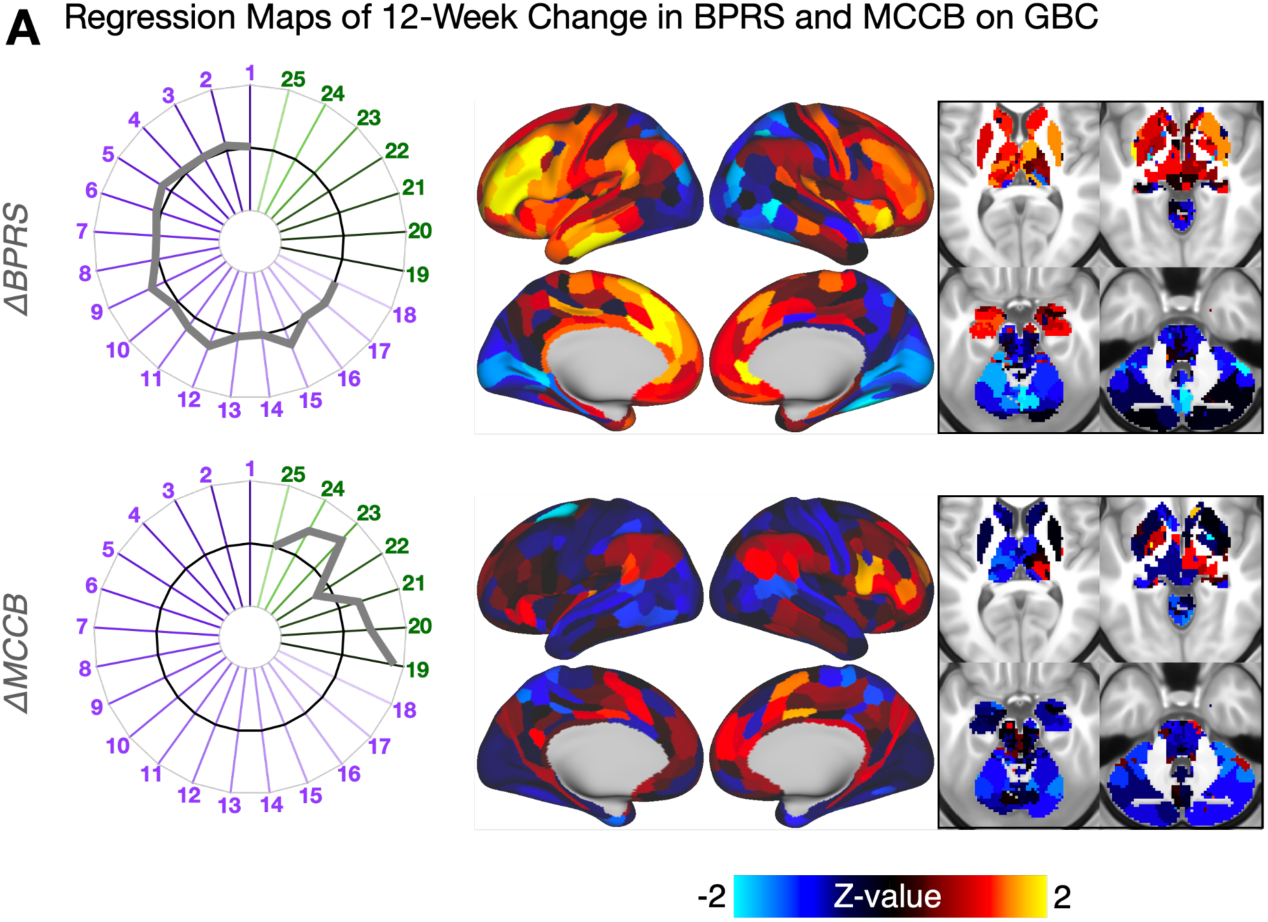
Characterization of the β_Δ_ maps. **A)** Mass univariate maps of the ΔBPRS and ΔMCCB scores onto GBC across 30 patients. Radarplots show the group mean change in total BPRS and MCCB scores over 12 weeks of clozapine treatment. Beta coefficient values in the neural maps are shown as Z-scores.

**Supplementary Figure 8.**
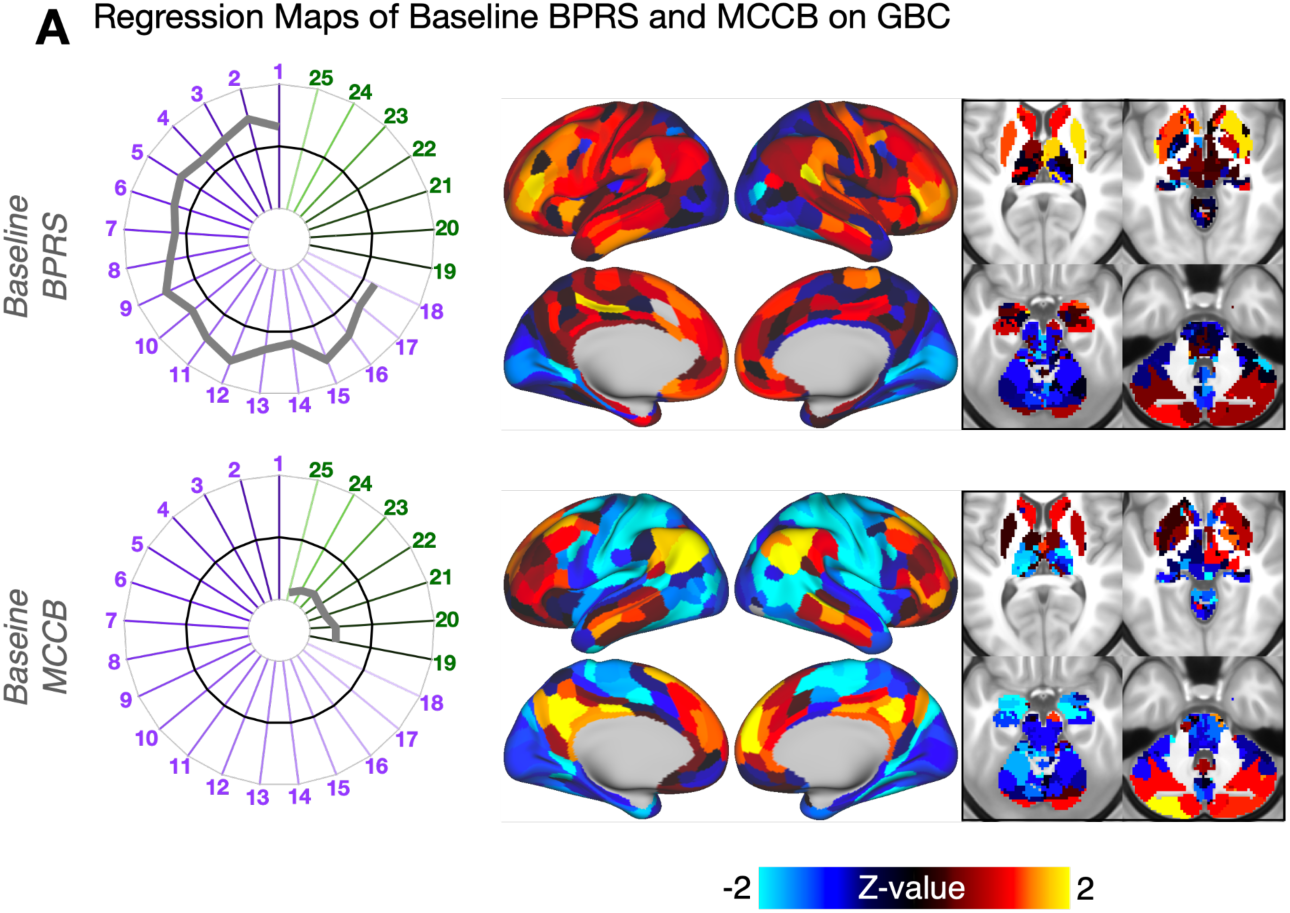
Characterization of the β_Week0_ maps. **A)** Mass univariate maps of the baseline BPRS and MCCB scores onto GBC across 30 patients. Radarplots show the group mean baseline total BPRS and MCCB scores. Beta coefficient values in the neural maps are shown as Z-scores.

**Supplementary Figure 9.**
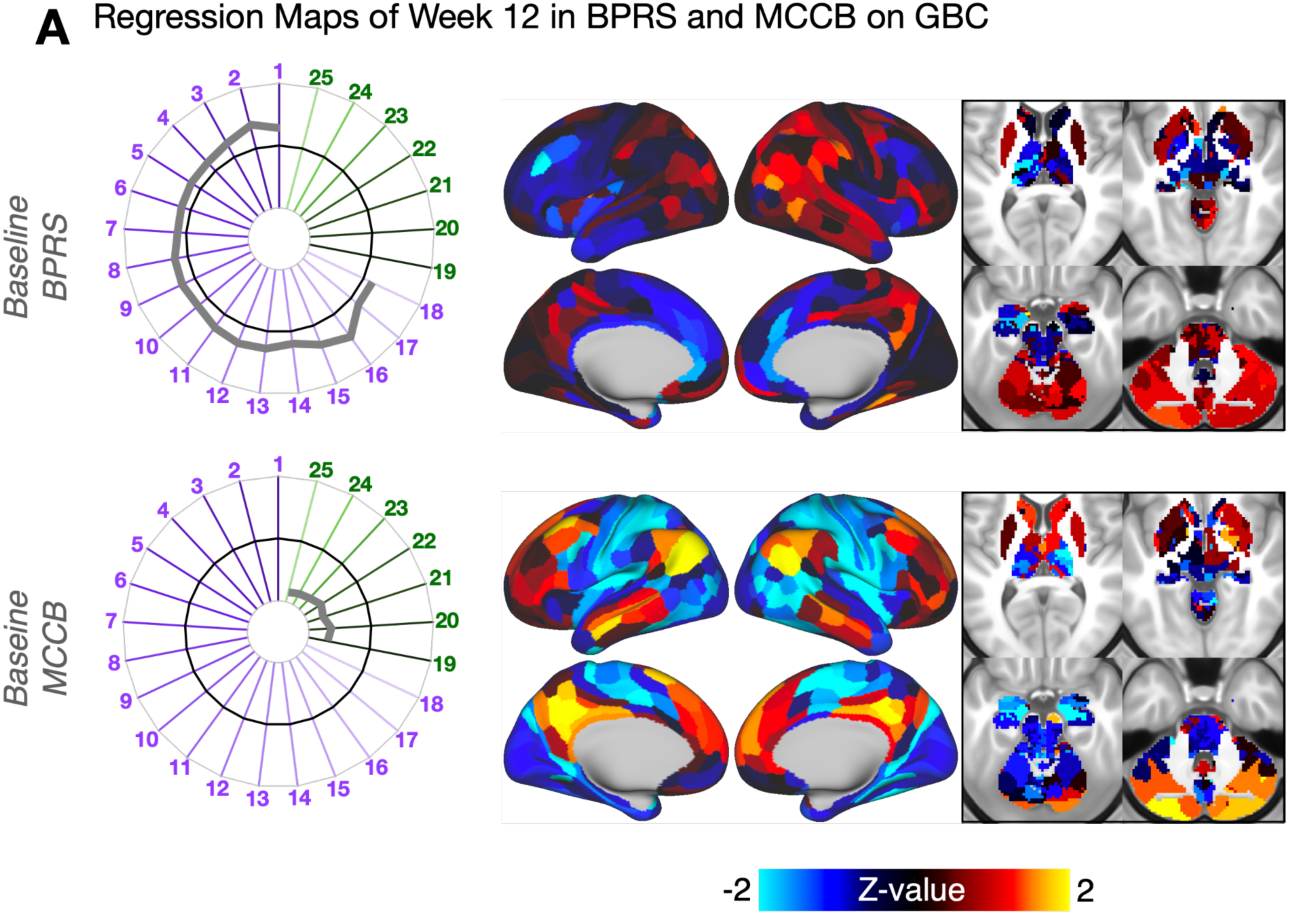
Characterization of the β_Week12_ maps. **A)** Mass univariate maps of the Week 12 residual BPRS and MCCB scores onto GBC across 30 patients. Radarplots show the group mean residual total BPRS and MCCB scores at Week 12. Beta coefficient values in the neural maps are shown as Z-scores.

**Supplementary Figure 10.**
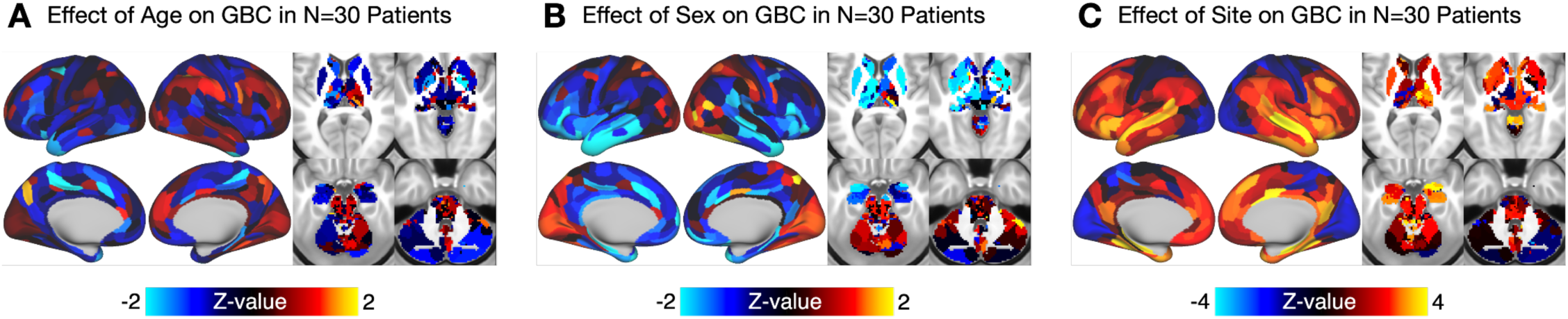
Effect of age, sex, and site on GBC across N=30 patients. **A)** Age at scan, **B)** sex (binarized as 1=male, 2=female), and **C)** site (1=ZHH, 2=CAMH) for each subject were included in each symptom-neural regression model as covariates of no interest, such that these variables would not drive the symptom-neural effects shown in the main text. Images shown are taken from the general linear model using PC1_Δ_ scores as the covariate of interest, but the maps of effects of age, sex, and site were highly similar across all models.

**Supplementary Figure 11.**
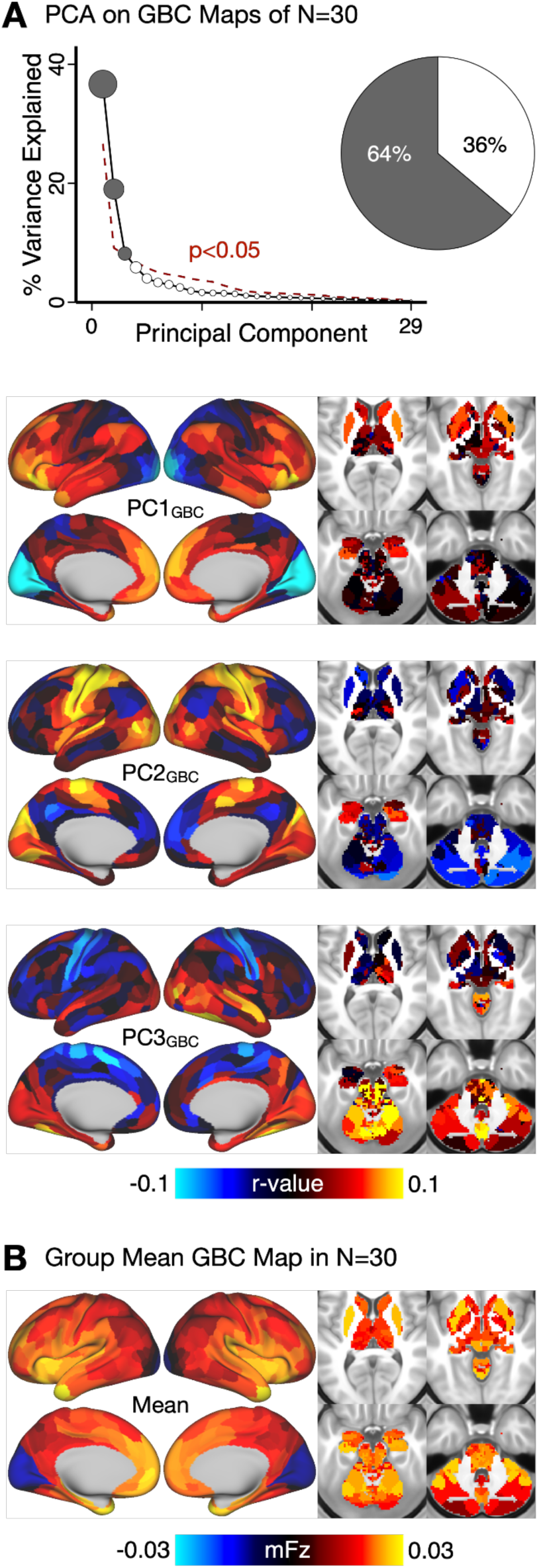
Principal components of GBC maps across patients. **A)** PCA performed on all neural GBC maps (PCA_GBC_). Permutation analysis shows that the first 3 components explain significantly more variance than expected by chance (screeplot). Together, they account for 64% of the total neural variance across patients (pie chart). The loadings on each neural parcel for the first 3 PCs are shown. The strength of the loading in each parcel reflects the contribution of that parcel to the neural variance captured by that PC across patients. **B)** For comparison, the group mean GBC map is also shown.

**Supplementary Figure 12.**
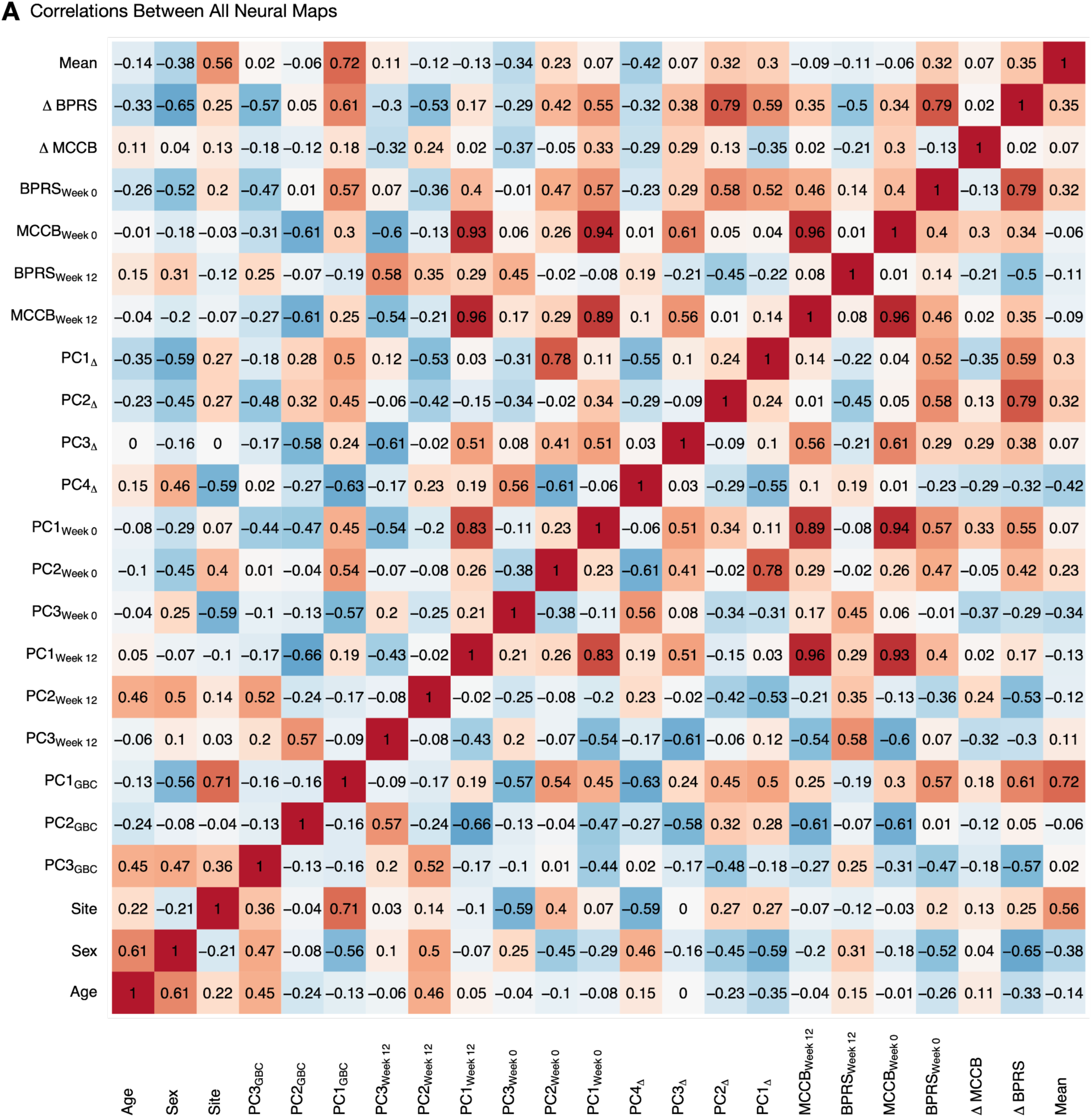
**A)** Correlations (Pearson’s r) between all neural maps. The map of PC1_GBC_ correlates strongly with the effect of Site as well as the group mean GBC map.

**Supplementary Table 1.** Demographics of the clinical sample.

| N=30 | Mean | SD |
| --- | --- | --- |
| Age at Baseline | 27.02 | 7.97 |
| Sex (% Male) | 53.33 | - |
| Baseline BPRS (Composite Score) | 43.33 | 9.18 |
| Baseline MCCB (Overall T-Score) | 25.27 | 13.10 |
| Week 12 BPRS (Composite Score) | 31.33 | 7.97 |
| Week 12 MCCB (Overall T-Score) | 27.83 | 14.70 |
| Max Clozapine Dosage (mg) | 255.96 | 80.40 |
| Modal Clozapine Dosage (mg) | 244.00 | 82.06 |
| Mean Clozapine Dosage (mg) | 194.47 | 66.58 |

**Supplementary Table 2.** Correlations (Pearson’s r) between gene expression maps of interest (derived as in (35)) and all neural target maps. Minimum and maximum surrogate r-values and p-values were computed between the gene expression maps and 100,000 surrogate maps, generated by shuffling the parcels while preserving the spatial autocorrelations of the target neural map (57). Significance is denoted by ** (α < 0.05, Bonferroni corrected across all comparisons).

| Gene | | $\beta_{PC1,\Delta}$ | | | | $\beta_{PC2,\Delta}$ | | | | $\beta_{PC3,\Delta}$ | | | | $\beta_{PC4,\Delta}$ | | | | Mean | | | | $\beta_{\Delta BPRS}$ | | | | $\beta_{\Delta MCCB}$ | | | |
| --- | --- | --- | --- | --- | --- | --- | --- | --- | --- | --- | --- | --- | --- | --- | --- | --- | --- | --- | --- | --- | --- | --- | --- | --- | --- | --- | --- | --- | --- |
| Name | Stability | r-value | min surr r | max surr r | p-value | r-value | min surr r | max surr r | p-value | r-value | min surr r | max surr r | p-value | r-value | min surr r | max surr r | p-value | r-value | min surr r | max surr r | p-value | r-value | min surr r | max surr r | p-value | r-value | min surr r | max surr r | p-value |
| HTR2A | 0.92 | 0.178 | -0.223 | 0.099 | 0.0E+00 ** | 0.400 | -0.246 | 0.069 | 0.0E+00 ** | -0.244 | -0.175 | 0.169 | 0.0E+00 ** | -0.231 | -0.115 | 0.196 | 0.0E+00 ** | 0.223 | -0.217 | 0.145 | 0.0E+00 ** | 0.294 | -0.234 | 0.091 | 0.0E+00 ** | -0.001 | -0.167 | 0.158 | 9.8E-01 |
| HTR2C | 0.80 | 0.174 | -0.149 | 0.167 | 0.0E+00 ** | 0.124 | -0.170 | 0.174 | 3.7E-03 | 0.078 | -0.182 | 0.176 | 9.6E-02 | -0.136 | -0.171 | 0.167 | 1.3E-03 | -0.048 | -0.198 | 0.198 | 3.7E-01 | 0.256 | -0.168 | 0.211 | 0.0E+00 ** | -0.070 | -0.181 | 0.172 | 8.9E-02 |
| DRD1 | 0.51 | 0.162 | -0.181 | 0.135 | 0.0E+00 ** | 0.225 | -0.191 | 0.116 | 0.0E+00 ** | -0.042 | -0.179 | 0.162 | 2.8E-01 | -0.212 | -0.145 | 0.180 | 0.0E+00 ** | 0.097 | -0.203 | 0.167 | 3.0E-02 | 0.259 | -0.192 | 0.146 | 0.0E+00 ** | -0.009 | -0.149 | 0.173 | 9.0E-01 |
| DRD2 | 0.89 | 0.094 | -0.130 | 0.203 | 8.4E-02 | -0.016 | -0.128 | 0.186 | 2.4E-01 | 0.151 | -0.177 | 0.194 | 3.6E-04 | -0.043 | -0.190 | 0.141 | 5.4E-01 | -0.177 | -0.207 | 0.197 | 4.0E-05 ** | 0.161 | -0.138 | 0.196 | 1.1E-03 | -0.091 | -0.163 | 0.170 | 2.5E-02 |
| DRD3 | 0.68 | 0.052 | -0.115 | 0.187 | 6.2E-01 | -0.092 | -0.108 | 0.186 | 6.0E-04 | 0.112 | -0.164 | 0.186 | 9.8E-03 | -0.006 | -0.188 | 0.141 | 6.6E-01 | -0.156 | -0.228 | 0.189 | 1.3E-03 | 0.022 | -0.136 | 0.194 | 7.1E-01 | -0.046 | -0.166 | 0.165 | 2.6E-01 |
| DRD5 | 0.82 | 0.226 | -0.194 | 0.121 | 0.0E+00 ** | 0.344 | -0.209 | 0.129 | 0.0E+00 ** | -0.145 | -0.181 | 0.178 | 4.4E-04 | -0.271 | -0.134 | 0.180 | 0.0E+00 ** | 0.147 | -0.188 | 0.182 | 5.6E-04 | 0.348 | -0.203 | 0.130 | 0.0E+00 ** | -0.066 | -0.154 | 0.188 | 9.2E-02 |
| HRH1 | 0.64 | 0.195 | -0.219 | 0.093 | 0.0E+00 ** | 0.417 | -0.244 | 0.080 | 0.0E+00 ** | -0.210 | -0.189 | 0.167 | 0.0E+00 ** | -0.232 | -0.115 | 0.198 | 0.0E+00 ** | 0.289 | -0.216 | 0.163 | 0.0E+00 ** | 0.348 | -0.233 | 0.107 | 0.0E+00 ** | -0.008 | -0.166 | 0.177 | 8.4E-01 |
| HRH2 | 0.51 | 0.181 | -0.190 | 0.129 | 0.0E+00 ** | 0.349 | -0.218 | 0.109 | 0.0E+00 ** | -0.217 | -0.174 | 0.165 | 0.0E+00 ** | -0.195 | -0.137 | 0.197 | 0.0E+00 ** | 0.122 | -0.210 | 0.170 | 2.9E-03 | 0.294 | -0.208 | 0.129 | 0.0E+00 ** | -0.080 | -0.164 | 0.159 | 3.9E-02 |
| HRH3 | 0.56 | -0.005 | -0.139 | 0.142 | 7.1E-01 | 0.114 | -0.111 | 0.205 | 1.5E-02 | 0.102 | -0.168 | 0.144 | 1.2E-03 | 0.002 | -0.169 | 0.152 | 6.7E-01 | 0.110 | -0.147 | 0.156 | 3.6E-03 | 0.215 | -0.131 | 0.182 | 0.0E+00 ** | -0.011 | -0.136 | 0.165 | 5.5E-01 |
| HRH4 | 0.13 | -0.044 | -0.111 | 0.216 | 1.1E-02 | -0.253 | -0.129 | 0.193 | 0.0E+00 ** | 0.087 | -0.141 | 0.190 | 8.1E-02 | 0.114 | -0.191 | 0.150 | 4.0E-04 | -0.228 | -0.151 | 0.193 | 0.0E+00 ** | -0.176 | -0.110 | 0.213 | 0.0E+00 ** | -0.099 | -0.166 | 0.156 | 2.1E-02 |
| CHRM1 | 0.55 | 0.194 | 0.206 | 0.101 | 0.0E+00 ** | 0.432 | -0.209 | 0.109 | 0.0E+00 ** | -0.211 | -0.174 | 0.174 | 0.0E+00 ** | -0.245 | -0.135 | 0.188 | 0.0E+00 ** | 0.255 | -0.202 | 0.166 | 0.0E+00 ** | 0.355 | -0.215 | 0.123 | 0.0E+00 ** | 0.000 | -0.158 | 0.178 | 9.4E-01 |
| CHRM2 | 0.29 | 0.129 | -0.169 | 0.131 | 0.0E+00 ** | 0.205 | -0.183 | 0.122 | 0.0E+00 ** | -0.120 | -0.160 | 0.164 | 2.1E-03 | -0.114 | -0.154 | 0.168 | 6.0E-04 | 0.020 | -0.168 | 0.169 | 5.4E-01 | 0.204 | -0.185 | 0.139 | 0.0E+00 ** | -0.120 | -0.152 | 0.154 | 9.0E-04 |
| CHRM3 | 0.91 | 0.201 | -0.206 | 0.108 | 0.0E+00 ** | 0.432 | -0.249 | 0.082 | 0.0E+00 ** | -0.240 | -0.194 | 0.165 | 0.0E+00 ** | -0.254 | -0.128 | 0.193 | 0.0E+00 ** | 0.251 | -0.218 | 0.155 | 0.0E+00 ** | 0.347 | -0.228 | 0.103 | 0.0E+00 ** | -0.025 | -0.169 | 0.171 | 5.0E-01 |
| CHRM4 | 0.21 | 0.088 | -0.124 | 0.178 | 9.4E-02 | 0.080 | -0.124 | 0.195 | 2.0E-01 | 0.079 | -0.167 | 0.197 | 5.5E-02 | -0.053 | -0.188 | 0.142 | 3.9E-01 | -0.095 | -0.168 | 0.184 | 2.4E-02 | 0.191 | -0.138 | 0.209 | 0.0E+00 ** | -0.048 | -0.171 | 0.161 | 2.3E-01 |
| CHRM5 | 0.58 | 0.084 | -0.166 | 0.173 | 6.3E-02 | -0.026 | -0.160 | 0.166 | 3.5E-01 | 0.021 | -0.155 | 0.169 | 7.1E-01 | -0.036 | -0.184 | 0.149 | 4.3E-01 | -0.186 | -0.173 | 0.150 | 0.0E+00 ** | 0.049 | -0.168 | 0.167 | 3.2E-01 | -0.092 | -0.155 | 0.165 | 1.5E-02 |
| ADRA1A | 0.13 | -0.031 | -0.151 | 0.158 | 2.9E-01 | -0.006 | -0.139 | 0.139 | 7.6E-01 | 0.061 | -0.157 | 0.163 | 1.4E-01 | 0.042 | -0.169 | 0.169 | 2.1E-01 | -0.046 | -0.176 | 0.158 | 2.6E-01 | -0.019 | -0.157 | 0.154 | 4.8E-01 | 0.039 | -0.154 | 0.147 | 2.5E-01 |
| ADRA1B | 0.79 | 0.123 | -0.212 | 0.097 | 0.0E+00 ** | 0.409 | -0.240 | 0.071 | 0.0E+00 ** | -0.229 | -0.176 | 0.150 | 0.0E+00 ** | -0.177 | -0.132 | 0.201 | 0.0E+00 ** | 0.333 | -0.227 | 0.169 | 0.0E+00 ** | 0.296 | -0.236 | 0.087 | 0.0E+00 ** | -0.024 | -0.161 | 0.167 | 4.8E-01 |
| ADRA1D | 0.33 | 0.233 | -0.186 | 0.144 | 0.0E+00 ** | 0.278 | -0.190 | 0.156 | 0.0E+00 ** | -0.178 | -0.190 | 0.181 | 4.0E-05 ** | -0.172 | -0.148 | 0.175 | 0.0E+00 ** | 0.015 | -0.206 | 0.199 | 6.8E-01 | 0.269 | -0.186 | 0.158 | 0.0E+00 ** | -0.128 | -0.189 | 0.180 | 1.9E-03 |
| ADRA2A | 0.28 | 0.104 | -0.179 | 0.132 | 2.6E-04 ** | 0.254 | -0.200 | 0.118 | 0.0E+00 ** | -0.197 | -0.148 | 0.170 | 0.0E+00 ** | -0.087 | -0.160 | 0.180 | 4.9E-03 | 0.198 | -0.185 | 0.171 | 0.0E+00 ** | 0.154 | -0.202 | 0.133 | 0.0E+00 ** | 0.025 | -0.171 | 0.155 | 4.3E-01 |
| ADRA2B | 0.45 | 0.004 | -0.122 | 0.190 | 2.6E-01 | -0.116 | -0.103 | 0.203 | 0.0E+00 ** | 0.177 | -0.152 | 0.164 | 0.0E+00 ** | 0.058 | -0.180 | 0.136 | 1.9E-02 | -0.150 | -0.160 | 0.185 | 8.0E-05 ** | 0.053 | -0.113 | 0.205 | 9.1E-01 | -0.076 | -0.151 | 0.164 | 4.4E-02 |
| ADRA2C | 0.84 | 0.120 | -0.170 | 0.154 | 2.6E-04 ** | 0.181 | -0.158 | 0.136 | 0.0E+00 ** | 0.072 | -0.151 | 0.155 | 6.9E-02 | -0.174 | -0.172 | 0.167 | 0.0E+00 ** | 0.120 | -0.183 | 0.154 | 2.2E-03 | 0.277 | -0.170 | 0.171 | 0.0E+00 ** | 0.007 | -0.170 | 0.154 | 8.1E-01 |

